# Disrupted topological properties of structural brain networks present a glutamatergic neuropathophysiology in people with narcolepsy

**DOI:** 10.1101/2023.07.25.23293138

**Authors:** Guoyan Chen, Wen Wang, Haoyang Wu, Xiangchao Zhao, Xiaopeng Kang, Jiafeng Ren, Jun Zhang, Jiaxiu He, Shihui Sun, Zhao Zhong, Danqing Shang, Mengmeng Fan, Jinxiang Cheng, Dan Zhang, Changjun Su, Jiaji Lin

**Affiliations:** Department of Neurology, The Second Affiliated Hospital of Air Force Medical University, No.569 Xinsi Road, Xi’an, 710038, China; Department of Radiology, The Second Affiliated Hospital of Air Force Medical University, No.569 Xinsi Road, Xi’an, 710038, China; Basic Medicine School, Air Force Medical University, No.169 Changle West Road, Xi’an, 710038, China; School of Artificial Intelligence, University of Chinese Academy of Sciences, No.19A Yuquan Road, Beijing, 100876, China; Department of Radiology, Chinese PLA General Hospital/Medical School of Chinese PLA, No.28 Fuxing Road, Beijing, 100853, China

**Author notes:** Correspondence to: Jiaji Lin, Department of Neurology, The Second Affiliated Hospital of Air Force Medical University, No.569 Xinsi Road, Xi’an, 710038, China. & Changjun Su, Department of Neurology, The Second Affiliated Hospital of Air Force Medical University, No.569 Xinsi Road, Xi’an, 710038, China. These authors contributed equally to this work (Guoyan Chen, Wen Wang, Haoyang Wu, Xiangchao Zhao).

**Keywords:** narcolepsy, structural network, topological property, glutamatergic receptor, cingulate gyrus

## Abstract

**Study objectives:** Growing evidences have documented various abnormalities of the white matter bundles in people with narcolepsy. We sought to evaluate topological properties of brain structural networks, and their association with symptoms and neuropathophysiological features in people with narcolepsy.

**Methods:** Diffusion tensor imaging (DTI) was conducted for people with narcolepsy (n = 30) and matched healthy controls as well as symptoms assessment. Structural connectivity for each participant was generated to analyze global and regional topological properties and their correlations with narcoleptic features. Further human brain transcriptome was extracted and spatially registered for connectivity vulnerability. Genetic functional enrichment analysis was performed and further clarified using *in vivo* emission computed tomography data.

**Results:** A wide and dramatic decrease in structural connectivities was observed in people with narcolepsy, with descending network degree and global efficiency. These metrics were not only correlated with sleep latency and awakening features, but also reflected alterations of sleep macrostructure in people with narcolepsy. Network-based statistics identified a small hyperenhanced subnetwork of cingulate gyrus that was closely related to rapid eye movement sleep behavior disorder (RBD) in narcolepsy. Further imaging genetics analysis suggested glutamatergic signatures were responsible for the preferential vulnerability of connectivity alterations in people with narcolepsy, while additional PET/SPECT data verified that structural alteration was significantly correlated with metabotropic glutamate receptor 5 (mGlutR5) and N-methyl-D-aspartate receptor (NMDA).

**Conclusions:** People with narcolepsy endured a remarkable decrease in the structural architecture, which was not only be closely related to narcolepsy symptoms but also glutamatergic signatures.

**Statement of Significance:** Growing evidences have identified a widespread disrupted white matter integrity of people with narcolepsy, so that connectome properties and neuropathophysiological features underlying these abnormalities have become a topic of increasing interest. This report extends on findings regarding the structural wirings and architectural topology of people with narcolepsy and inferring their clinical correlation with sleepiness assessment, polysomnography features and sleep macrostructure. Further imaging genetics analysis suggests glutamatergic signatures are responsible for the preferential vulnerability of connectivity alterations, while additional PET/SPECT data verifies that structural alteration is significantly correlated with metabotropic glutamate receptor 5 (mGlutR5) and N-methyl-D-aspartate receptor (NMDA). Our findings, therefore, converge structural network and genetic signatures for in people with narcolepsy.

## 1. Introduction

Narcolepsy is conceptualized as a state of instability or loss of boundary control that manifests as an inability to remain in sleep or wake states for the normal length of time and by the inappropriate occurrence of sleep phenomena during wakefulness, and vice versa ^1^. Its onset peaks in the second decade of life and affects 1 in 2,000 people, and most people with narcolepsy also have symptoms indicative of cataplexy, sleep paralysis, hypnagogic hallucinations and disturbed nocturnal sleep ^1^. A variety of neuroplastic alterations have been found to be closely associated with this wakefulness/sleep disturbances. Positron emission tomography (PET) study of Trotti et al. showed hypermetabolism in precuneus, inferior parietal lobule, superior and middle temporal gyri, and culmen ^2^. In another study that used functional magnetic resonance imaging (fMRI), functional connectivity was decreased between regions of the limbic system and the default mode network (DMN) and increased in the visual network ^3^. Furthermore, N-acetylaspartate/creatine-phosphocreatine in hypothalamus and myo-Inositol/ creatine-phosphocreatine in the amygdalain was reported to link with narcolepsy in 1H-MRS study of Joo et al ^4^. Although disrupted functional activities have been widely reported in narcolepsy, the potential clinical implications of structural connectome abnormalities have not been systematically investigated.

The key attribute of the structural connectome is the large investment of neural resources in macroscale connectivity to keep the brain globally connected, with a considerable proportion of the anatomical communication pathways comprising white matter^5^. Cross-species studies on these wirings have revealed common “principles of network” appear to be widely conserved and represent fundamental features of brain organization and function ^6^. Considerable effort in the past decade has been directed towards identifying network substrates and biomarkers that predict disease severity, prognosis and outcome of specific brain disorders, such as deglobalization in the DMN and motor network in Alzheimer disease and ALS ^7,8^, fluctuation in functional network dynamics in epilepsy ^9^, hyperconnectivity in autism spectrum disorder ^10^, and so on. In recent years, growing evidences from diffusion tensor imaging (DTI) have documented a number of abnormalities of the white matter bundles in people with narcolepsy. For example, Gool et al. have found brain-wide significantly lower fractional anisotropy (FA) in sleep-wake regulation-related, limbic and reward system areas of drug-free people with narcolepsy type 1, which was sensitive to the fibers alignment and the myelination integrity ^11^. Similar FA decreases have also been confirmed in the bilateral cerebellar hemispheres, bilateral thalami, corpus callosum and midbrain white matter of people with narcolepsy type 2 ^12,13^. Therefore, further analysis with network neuroscience tools may help us to discover topological features specific to this disease.

Therefore, current study obtained MR imaging from people with narcolepsy for structural connectome analysis. By characterizing alteration of global and regional signatures, we further associated them to clinical symptoms as well as underlying neuropathophysiological susceptibilities, which was further clarified by *in vivo* emission computed tomography data.

## 2. Materials and methods

### 2.1 Experiment design and participants

This was a prospective observational neuroimaging study aimed at exploring the clinical and brain imaging features of people with narcolepsy (NCT03376568), which was registered in 2016. It was approved by the Ethics Committee of the Second Affiliated Hospital (also named as Tangdu Hospital) of Air Force Medical University, and all participants and their families were fully informed and signed an informed consent form for the operation. From December 2017 to December 2022, over 100 participants complaining about daily periods of irrepressible need to sleep or daytime lapses into sleep occurring for at least three months were recruited at our narcolepsy clinic and signed up for the study. All subjects had to be 18-65 years old, treatment-naïve, right-handed according to the Edinburgh Handedness Scale ^14^ and have normal or corrected-to-normal vision. To be included, people should be diagnosed as narcolepsy type 1/2 (with/without cataplexy) according to the 3rd edition of the International Classification of Sleep Disorders (ICSD-3) ^15^. Briefly, they took sleep diaries and actigraphy for two weeks for exclusion of sleep deprivation, insufficient sleep syndrome and circadian rhythm sleep-wake disorders. Any medication or intervention should be avoided during this period, and sleep breathing disorder, depression or other potential sleep disorders were also excluded. At the end, polysomnography (PSG) and MSLT were performed according to standard techniques as AASM recommendation ^16^. A mean sleep latency of ≤ 8 minutes and two or more sleep onset REM periods (SOREMPs) on multiple sleep latency test (MSLT) performed according to standard techniques. A SOREMP (within 15 minutes of sleep onset) on the preceding nocturnal PSG may replace one of the SOREMPs in the MSLT. All recruited participants received MRI acquisition and symptom assessment on the next day.

The exclusion criteria included severe systematic diseases, central neurodegenerative diseases, cognitive impairment, unstable psychiatric diseases, structural brain abnormalities, history of intracranial hemorrhage or strokes, history of bleeding or coagulation abnormalities, inability to tolerate prolonged stationary position when supine, history of brain surgery, and contraindications for MRI scanning. Beck depression/anxiety inventory was used to initially screen out psychiatric disorder (cutoff ≤ 4/13 respectively), while Mini-Mental Status Exam (MMSE) was the tool used to screen out obvious cognitive dysfunction (cutoff = 27, adjusted by education level as need). The data of 30 participants with narcolepsy were finally included for present study, and additional 30 healthy controls (HCs) with matched gender, age and education level were also enrolled by the same exclusion criteria at the same time and received the same MRI scanning as people with narcolepsy. All raw data are available on request within the conditions of our ethics approval.

### 2.2 Narcoleptic symptom assessment

In addition to general characteristics (age, gender and previous medication use), participants and their first-degree relatives underwent clinical consultations and semi-structured interviews about narcoleptic symptoms (excessive daytime sleepiness (EDS), rapid eye movement (REM) sleep behavior disorder (RBD), cataplexy, sleep paralysis, and hypnagogic hallucination). The Epworth Sleepiness Scale (ESS) (Johns, 1991) was administered as a measure of EDS. ESS score > 10 is defined as EDS. The diagnose of RBD is were characterized by any complaining of probable RBD (repeated sleep related vocalization and/or complex motor behaviors) and REM sleep without atonia (RWA) with the guidance of AASM manual v2.4 ^16^.

We also ascertained a wide range of PSG parameters, including total sleep time (TST), sleep efficiency with respect to both total registration and total sleep time, percentage of sleep stages N1, N2, N3, and REM (with respect to total sleep time), sleep latency to the first occurrence of any sleep stage from light off, wake time after sleep onset (WASO), awakening number per hour (AWN) and apnea-hypopnea index (AHI). Technicians awakened the participants after a registration time of roughly 7.5 hours.

### 2.3 MR imaging data acquisition

All MR images were acquired using a 3.0T MR imaging system (Discovery 750; GE Healthcare, Milwaukee, WI, USA) equipped with a 64-channel phase array coil. Both Diffusion tensor imaging (DTI) and High-resolution sagittal 3D T1-weighted imaging (3D T1WI) were operated under quality control and assurance guidelines in accordance with the recommendations of the American College of Radiology and utilized the same imaging sequence. Participants were instructed to lie supine, relax, remain still, keep awake with eyes closed, and think of nothing during MRI acquisition. They wore ear plugs to reduce noise. To minimize head motion, tight but comfortable foam padding was placed around their heads. The MRI scanning protocol included the following: (1) DTI: repetition time/echo time (TR / TE) = 7,522/80.8 ms; field of view (FOV) = 224 × 224; matrix = 112 × 112; b value = 1,000 sec/mm^2^; encoding directions = 64; slice thickness = 2 mm; and B_0_ = 4. (2) 3D T1WI: TR/TE = 6.656 / 2.928 ms; inversion time = 800 ms; FA = 7°; FOV = 256 mm × 256 mm; matrix = 256 × 256; slice thickness = 1 mm; contiguous slices = 190. Two senior radiologists independently reviewed the aforementioned sequences, and carefully checked the quality of MRI images.

### 2.4 Network construction and subnetwork extract

DTI data were preprocessed using the DiffusionKit toolbox as previous report ^17^. Briefly, the process included: (1) converting to DICOM data; (2) extracting a brain mask; (3) correcting for head motion and eddy current; (4) fitting diffusion tensors and performing tractography with a deterministic streamline method with the fractional anisotropy (FA) threshold of 0.3 and the angular threshold of 45°; (5) registering regions of interest (ROIs) from Montreal Neurological Institute (MNI) space to the individual space; (6) pruning tract for each ROI pair in the Brainnetome atlas (atlas.brainnetome.org) ^18^. Brainnetome atlas includes middle frontal gyrus (MFG), parahippocampal gyrus (PhG), cingulate gyrus (CG), basal ganglia (BG) and other contralateral 24 partitions. The streamline number was referred to as connectivity by calculating for the tracts linking each pair of brain regions. All connectivity pairs were collected and then combined into 246□×□246 undirected and weighted connectivity matrix as global structural network. All ROI full names and abbreviations of the Brainnetome atlas could be found in Supplementary Table 1.

Considering the regional variation in the structural subnetwork, network-based statistic was used to identify expansive disconnected subnetworks ^19^. This non-parametric connectome-wide method is based on the principles underpinning traditional cluster-based thresholding of statistical parametric maps, and exploits the extent to which the connections comprising the contrast or effect of interest are interconnected ^19^. Network-based statistics were performed with 5,000 permutations for weighted structural network comparisons between the participants and matched HCs. A threshold of *t*□=□3 were applied for streamline comparison with a significance of *P*□<□0.05 for permutation test.

### 2.5 Graph-Theory network analysis

We applied a sparsity threshold of 10% for the weighted network of each participant to avoid false connections. All graph-theory network analyses in this study were performed using the Brant toolbox (https://github.com/yongliulab/brant/) ^20^. Topological metrics were calculated for each participant, reflecting different aspects of the structural network properties, including: (1) network/nodal degree is the most commonly used as a measure of the wiring strength and coherence of anatomical network/ROI. The degree of an individual node not only reflects the cumulative “wiring cost” distribution of the regional network, but is also used as a measure of density distribution ^21^. (2) global/nodal efficiency is measure of integration, which assessed parallel information transfer within a structural network/ROI. (3) clustering coefficient is measure of segregation, which is known as the fraction of triangles around an individual node, and the mean clustering coefficient for the network reflects, on average, the prevalence of clustered connectivity. Detailed calculation formulas of these metrics are the same as described in the works of Sporns et al ^21–25^.

### 2.6 Spatial alignment to neuropathophysiological features

We investigated the underlying neuropathophysiological features of people with narcolepsy using the human brain gene transcriptome data extracted from the Allen Human Brain Atlas (AHBA) ^26^. This atlas is the first anatomically and genomically comprehensive three-dimensional human brain map, which contains a database of 20,737 gene expression levels represented by 58,692 probes. We matched the gene expression for each ROI to the Brainnetome atlas using the Abagen toolbox ^27,28^, by setting the threshold for intensity-based filtering of probes to 0.5. In the end, 236 ROI of 15,633 genes were extracted, resulting in a 236 × 15,633 matrix for the alteration of nodal degree. Partial least squares (PLS) regression algorithm, the statistically inspired modification of the partial least squares (SIMPLS), was applied to investigate how genetic variance can explain brain structural alterations ^29^. The ranked gene list with principle PLS weights was fed into the online tool WebGestalt ^30^ to identify the functional enrichment by gene set enrichment analysis (GSEA) ^31^. The false discovery rate (FDR) correction was applied, along with a significance of *P*_FDR_ <□0.05 for all enrichment analyses. The entire workflow is shown in Supplementary Figure 1.

To further elucidate our GSEA results, we also introduced PET/SPECT maps of glutamatergic neuropathophysiological features from unrelated control groups for spatial alignment, which were collected by Justine et al ^32^. Specifically, the [^11^C]ABP688 data for the group I metabotropic glutamate receptor 5 (mGluR5) expression came from Rosa-Neto team, Dubois team and Smart team respectively ^32–34^, while [^18^F]GE-179 data for the ionotropic N-methyl-D-aspartate receptor (NMDA) expression came from Galovic’s work ^35^. The correlation was calculated between the effect sizes of the ROI *t*-values and regional sum PET/SPECT values, resulting in a single measure for accessing the relationship between the altered brain structure and the glutamatergic features. To avoid false-positive results of spatial autocorrelation (SA), we generated 5,000 SA-preserving surrogate maps as null models using brainSMASH as null models ^36^. The significance was calculated as 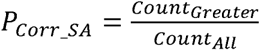 where *Count_Greater_* was the number that surrogate’s absolute correlation coefficient was greater than that of the primary analysis. *Count_All_* were the times that performed surrogate correlation, in this case, 5,000 times.

### 2.7 Statistical Analysis and code availability

Chi-square tests were used to detect group differences between people with narcolepsy and matched HCs in the demographic analysis. Non-paired *t*-tests were used evaluate the difference in each connectivity between people with narcolepsy and matched HCs, as well as graph metrics comparisons between narcolepsy and HCs group in the graph-theory network analysis. We further divided the people with narcolepsy into subgroups according to their clinical symptoms (cataplexy, sleep paralysis, hypnagogic hallucination, and RBD), and compared their graph metrics using non-paired *t*-tests too. Multiple comparison correlations of FDR were applied in both network and regional topology features (marked with *P*_FDR_). When analyzing clinical relevance, Pearson and Spearman correlation coefficient served as a measure. In order to screen out the best items that fit the predictive model, we adjusted the significant value by using the Bonferroni correction (marked with *P*_Bonferroni_). So that predictive performance of network degree was evaluated for MSLT sleep latency by stepwise linear models with leave-one-out cross-validation (LOOSWR). Network degrees were fit into linear regression with adjustment of subject’s sex, age, and constant. Using both forward and backward iterations (entered a model), iterations with a significance < 0.05 were count and its proportion was analyzed. Predicted MSLT sleep latency were compared using Pearson correlation coefficients too. For all the statistical analysis, the level of significance was set at *P* < 0.05. Majority of the computations were performed in the Python engine. All the statistical process and analysis code are available at https://github.com/Jiaji-Lab.

## 3. Results

### 3.1 Basic characteristics of people with narcolepsy

The data of 30 participants with narcolepsy (Male: Female = 18: 12) were finally collected, of which 56.67% (17/30) were type 1 with cataplexy and 43.33% (13/30) were type 2 without cataplexy. The mean age (± SD) of all people with narcolepsy was 29.5 ± 13.5 years. People with narcolepsy explicitly complained of EDS with a mean duration of 10.0 ± 7.0 years, with significantly higher ESS score than matched HCs (15.4 ±5.2 *vs.* 2.8 ± 2.0, *P* < 0.001). Moreover, 20.00% (6/30) of people with narcolepsy had sleep paralysis, 40.00% (12/30) hypnagogic hallucinations, and 63.33% (19/30) RBD, respectively. In the present study, all people with narcolepsy are treatment naïve and no other chronic medication has been found. There were 76.67% (23/30) of people with narcolepsy had nSOREMP in diagnostic PSG examination. It was also found that all the people with narcolepsy had more than 2 SOREMPs in MSLT test with low MSLT sleep latency of 2.25 ± 1.39 min. Brain MRI revealed no gross abnormalities in any participants with narcolepsy or HCs. Detail demographic and narcoleptic characteristics were summarized in Table 1.

**Table 1.** Characteristics of the study population.

|  | **Patients**<br>**(n = 30)** | **Matched HCs**<br>**(n = 30)** | **P value** |
| --- | --- | --- | --- |
| **Basic information** |  |  |  |
| Male: Female | 18:12 | 18:12 | $P > 0.05$ |
| Age (years, mean, SD) | 29.5 (13.5) | 29 (12.8) | $P > 0.05$ |
| **Narcolepsy features** |  |  |  |
| NT1/NT2 (n/n) | 17/13 | - | - |
| EDS (n/total) | 30/30 | 0/30 | $P < 0.001$ |
| EDS duration (years, mean, SD) | 10.0 (7.0) | - | - |
| ESS score (mean, SD) | 15.4 (5.2) | 2.8 (2.0) | $P < 0.001$ |
| Cataplexy (n/total) | 17/30 | 0/30 | $P < 0.001$ |
| Sleep paralysis (n/total) | 6/30 | 0/30 | $P < 0.001$ |
| Hypnagogic hallucinations (n/total) | 12/30 | 0/30 | $P < 0.001$ |
| RBD (n/total) | 19/30 | 0/30 | $P < 0.001$ |
| RBD in NT1 (n/n) | 12/17 | - | - |
| RBD in NT2 (n/n) | 7/13 | - | - |
| **Night polysomnography and MSLT** |  |  |  |
| TST (minutes, mean, SD) | 411.7 (53.4) | - | - |
| Sleep efficiency (minutes, mean, SD) | 85.6 (10.6) | - | - |
| Sleep latency (minutes, mean, SD) | 9.0 (8.7) | - | - |
| REM latency (minutes, mean, SD) | 88.9 (77.8) | - | - |
| nSOREMP (n/total) | 23/30 | - | - |
| N1% (mean, SD) | 29.3 (10.3) | - | - |
| N2% (mean, SD) | 39.5 (12.6) | - | - |
| N3% (mean, SD) | 13.8 (6.4) | - | - |
| REM times (mean, SD) | 4.6 (1.3) | - | - |
| REM% (mean, SD) | 17.5 (6.2) | - | - |
| Wake time after sleep onset (WASO) (mean, SD) | 26.5 (24) | - | - |
| Awakening number (AWN) per hour (mean, SD) | 4.0 (3.4) | - | - |
| Arousal index (AI) (mean, SD) | 3.8 (5.7) | - | - |
| Apnea-hypopnea index (AHI) (mean, SD) | 2.6 (2.3) | - | - |
| Limb movement (LM) index (mean, SD) | 22.1 (18.6) | - | - |
| Periodic leg movements (PLM) index (mean, SD) | 9.1 (16.9) | - | - |
| Mean sleep latency in MSLT (minutes, mean, SD) | 2.25 (1.39) | - | - |
| SOREMPs in MSLT (mean, SD) | 4.31 (0.99) | - | - |

Note: HC: health control; NT1: narcolepsy type 1; NT2: narcolepsy type 2; EDS: excessive daytime sleepiness; RBD: rapid eye movement (REM) sleep behavior disorder; ESS: Epworth Sleepiness Scale; MSLT: multiple sleep latency test; TST: total sleep time; nSOREMP = nocturnal SOREMP in PSG; SOREMP = the sleep-onset REM period.

### 3.2 Topology features of structural network in people with narcolepsy

To study the alterations in brain structural network of people with narcolepsy, all the structural connectivities of each participant were constructed. Non-paired *t*-test for each structural connectivity was calculated between people with narcolepsy and matched HCs, and all *t*-values were collected for global contrast matrix and converted into *t*-values distribution (Fig. 1A). As *t*-values distribution shown, a wide range of structural connectivities were found to endure a remarkable decrease without FDR correction in people with narcolepsy compared with matched HCs, as well as a small fraction of enhanced connectivities (Fig. 1B and Supplementary Figure 2). The most significant decreased structural connectivity were right ventrolateral area 6 (A6vl_R) of MFG to right inferior frontal junction (IFJ_R) of MFG (*t* = −5.594, *n* = 30), left caudal hippocampus (cHipp_L) to left caudal temporal thalamus (cTtha_L) (*t* = −5.481, *n* = 30), and left rostral area 35/36 (A35/36r_L) of PhG to cHipp_L (*t* = −5.328, *n* = 30).

**Figure 1.**
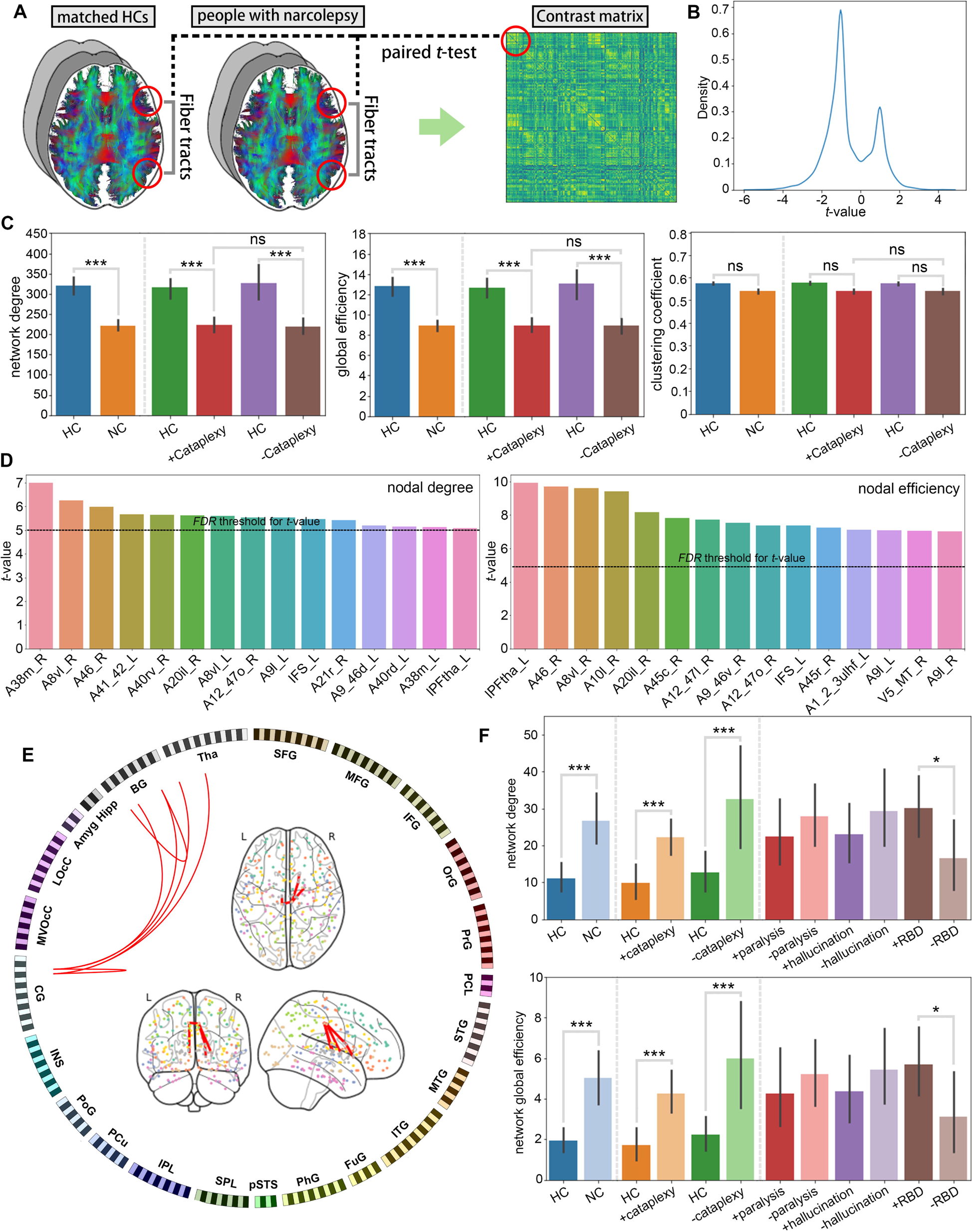
Topology features of structural network in people with narcolepsy. (**A**) An overview for building the contrast matrix. (**B**) The narcolepsy featured *t*-values distribution. (**C**) Group comparison of topological features in the global structural network of people with narcolepsy and matched HCs as well as subgroups. (**D**) The *t*-value in the comparison of regional topological features (nodal degree and nodal efficiency), where the dashed line represented the *t*-value threshold after FDR correction. (**E**) Network-based statistics revealed a subnetwork of hyperenhanced connectivity strength in people with narcolepsy. **(F)** Group comparison of topological features in the RBD-sensitive subnetwork of people with narcolepsy and matched HCs as well as subgroups. (^ns^*P*_FDR_ > 0.05, \**P*_FDR_ < 0.05, \*\*\**P*_FDR_ < 0.001)

Graph metrics of global structural network were calculated and a significant decrease in the network degree in people with narcolepsy were observed compared to matched HCs (*P*_FDR_ < 0.001) as well as global efficiency (*P*_FDR_ < 0.001, Fig. 1C). There was no significant difference in clustering coefficient between narcolepsy and HC group (Fig. 1C). We further divided all participants with narcolepsy into +cataplexy group (narcolepsy type 1, n = 17) and -cataplexy group (narcolepsy type 2, n = 13). Although both network degree and global efficiency showed a significant decline in +/−cataplexy participants compared with their matched HCs (network degree: *P*_FDR_ < 0.001; global efficiency: *P*_FDR_ < 0.001; Fig. 1C), there was no significant difference in the comparison between +cataplexy group and -cataplexy group (*P*_FDR_ > 0.05; Fig. 1C). Moreover, no significant difference was found for other clinical characteristics (sleep paralysis, hypnagogic hallucination and RBD; data not shown) among people with narcolepsy. For regional metrics analysis, nodal degree of 33 ROIs and nodal efficiency of 150 ROIs were found endured significant declines after FDR correction of all ROIs (*P*_FDR_ < 0.05, Fig. 1D). Further detailed information could be found in the Supplementary Table 2-3.

As for small but significant positive peak in *t*-values distribution of contrast matrix, we tried network-based statistics to better identify the hyperenhanced subnetwork. The analysis revealed a small subnetwork within 7 ROIs (*P*_FDR_□<□0.001), including both sides of caudal area 23 (A23c_L and A23c_R) of CG, right ventral caudate (vCa_R), globus pallidus (GP_R) and dorsal caudate (dCa_R) of BG, and left medial pre-frontal thalamus (mPFtha_L) and rostral temporal thalamus (rTtha_L) of Tha (Fig. 1E). Graph metrics of this subnetwork were not only found increased in the people with narcolepsy (network degree: *P*_FDR_ <□0.001; global efficiency: *P*_FDR_□<□0.001, Fig. 1F), but also presented a similar significant increase in +/−cataplexy participants compared with their matched HCs (network degree: *P*_FDR_ < 0.001; global efficiency: *P*_FDR_ < 0.001; Fig. 1F). Although no significant alteration could be found in either +/−paralysis or +/−hallucination comparison, there was also a significant increased in the people with narcolepsy with RBD *vs.* without RBD (network degree: *P*_FDR_□<□0.05; global efficiency: *P*_FDR_□<□0.05; Fig. 1F). So that it was referred to as the RBD-sensitive subnetwork.

### 3.3 Linking structural topology to clinical characteristics of people with narcolepsy

We analyzed the correlation between graph metrics and clinical characteristics within people with narcolepsy. Although neither network degree nor global efficiency was correlated with gender, age, ESS score, or REM latency in PSG, it turned out that both network degree and global efficiency were positively correlated with MSLT sleep latency (network degree: *r* = 0.59, *P* < 0.001; global efficiency: *r* = 0.55, *P* = 0.002; Fig.2A). Similarly, they were also positively correlated with sleep latency in PSG (network degree: *r* = 0.54, *P* = 0.002; global efficiency: *r* = 0.51, *P* = 0.004; Fig.2A). Moreover, they endured a significant negative correlation with WASO (network degree: *r* = −0.45, *P* = 0.013; global efficiency: *r* = −0.45, *P* = 0.012) and AWN (network degree: *r* = −0.38, *P* = 0.039; Fig.2B). As for sleep macrostructure, global efficiency but not network degree was negatively correlated with the N1 sleep time (*r* = −0.36, *P*L*=*L0.047) and percentage (*r* = 0.37, *P*L*=*L0.043; Fig.2C). Not only the network degree was positively correlated with the N2 sleep time (*r* = 0.44, *P*L*=*L0.015) and percentage (*r* = 0.38, *P*L*=*L0.037), but also the network global efficiency was significantly correlated (N2 sleep time: *r* = 0.51, *P*L*=*L0.004; N2 sleep percentage: *r* = 0.46, *P*L*=*L0.009; Fig.2C). No metrics was found correlated with N3 or REM sleep or other clinical characteristics. Finally, we also studied the graph metrics of RBD-sensitive subnetwork, and it only had significant correlation with RBD symptoms (subnetwork degree: spearman *r* = 0.36, *P*L*=*L0.0482; subnetwork global efficiency: spearman *r* = 0.41, *P*L*=*L0.024). No other significant correlation was found for RBD-sensitive subnetwork within people with narcolepsy.

**Figure 2.**
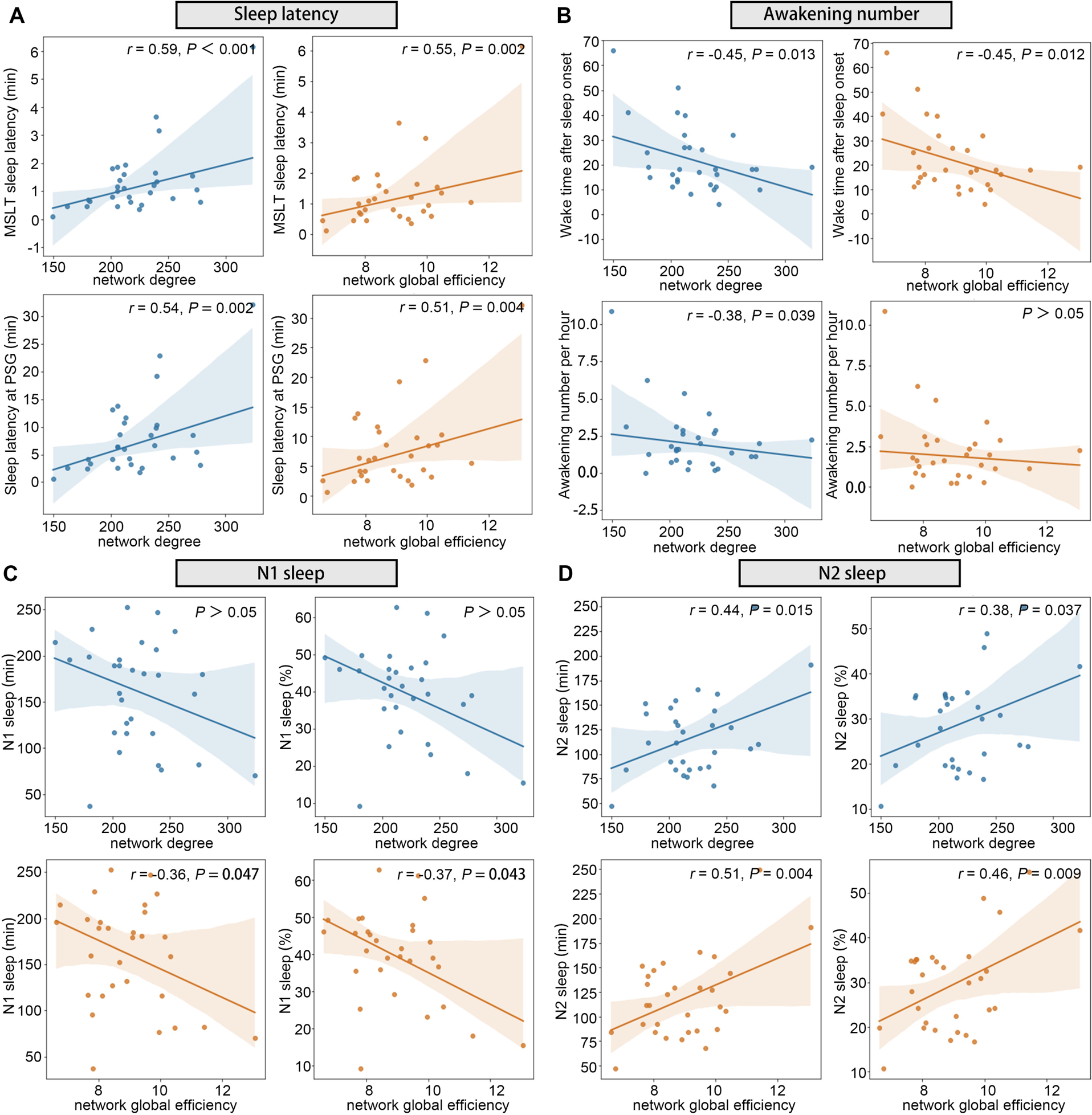
Linking structural topology to clinical characteristics of people with narcolepsy. Correlation results between metrics of global structural network of people with narcolepsy and MSLT sleep latency (**A**), sleep latency at PSG (**A**), wake time after sleep onset (**B**), awakening number per hour (**B**), N1 sleep time and percentage (**C**) and N2 sleep time and percentage (**D**).

We adjusted the significant value of clinical correlation by using the Bonferroni correction. As remarkable correlation with MSLT sleep latency (*P*_Bonferroni_ < 0.05), network degree was also applied in LOOSWR for MSLT Sleep latency prediction with adjustment of gender and age. It was found that network degree could be used as reliable biomarkers for MSLT sleep latency prediction (*r* = 0.71; iterative repetition = 100%, *P*_FDR_ < 0.05).

### 3.4 Linking structural narcolepsy topology to glutamatergic neuropathophysiology

We performed a PLS-based gene analysis to identify genes that were highly correlated with alterations of structural network in people with narcolepsy. Gene expression networks of both hemispheres of the healthy adult brain were constructed using the AHBA ^37^. The primary PLS features of narcolepsy genes accounted for 21.19% of the total variance with the *P* < 0.001 based on 5,000 permutations. These rank-list of narcolepsy-featured genes were annotated, and the Panther pathway analysis identified that these featured genes were functionally enriched in Metabotropic glutamate receptor (mGlutR) group I pathway (P00041, *P*_FDR_ <□0.001), Ionotropic glutamate receptor (iGlutR) pathway (P00037, *P*_FDR_ <□0.001) and Interferon-gamma (IFNγ) signaling pathway (P00035, *P*_FDR_ <□0.001; Fig. 3A). We then further investigated their GO characteristics. A post-processing step of affinity propagation revealed that the representative biological processes of the narcolepsy-featured genes were synaptic vesicle cycle (GO0099504), regulation of synapse structure or activity (GO:0050803), glutamate receptor signaling pathway (GO0007215) and regulation of membrane potential (GO0042391) (all *P*_FDR_ <□0.001; Fig. 3B). Majority of cellular components focused on synaptic membrane (GO: 0097060) and glutamatergic synapse (GO:0098978) (all *P*_FDR_ <□0.001; Fig. 3B). All the top gene functional properties were provided in Supplementary Tables 4-6.

**Figure 3.**
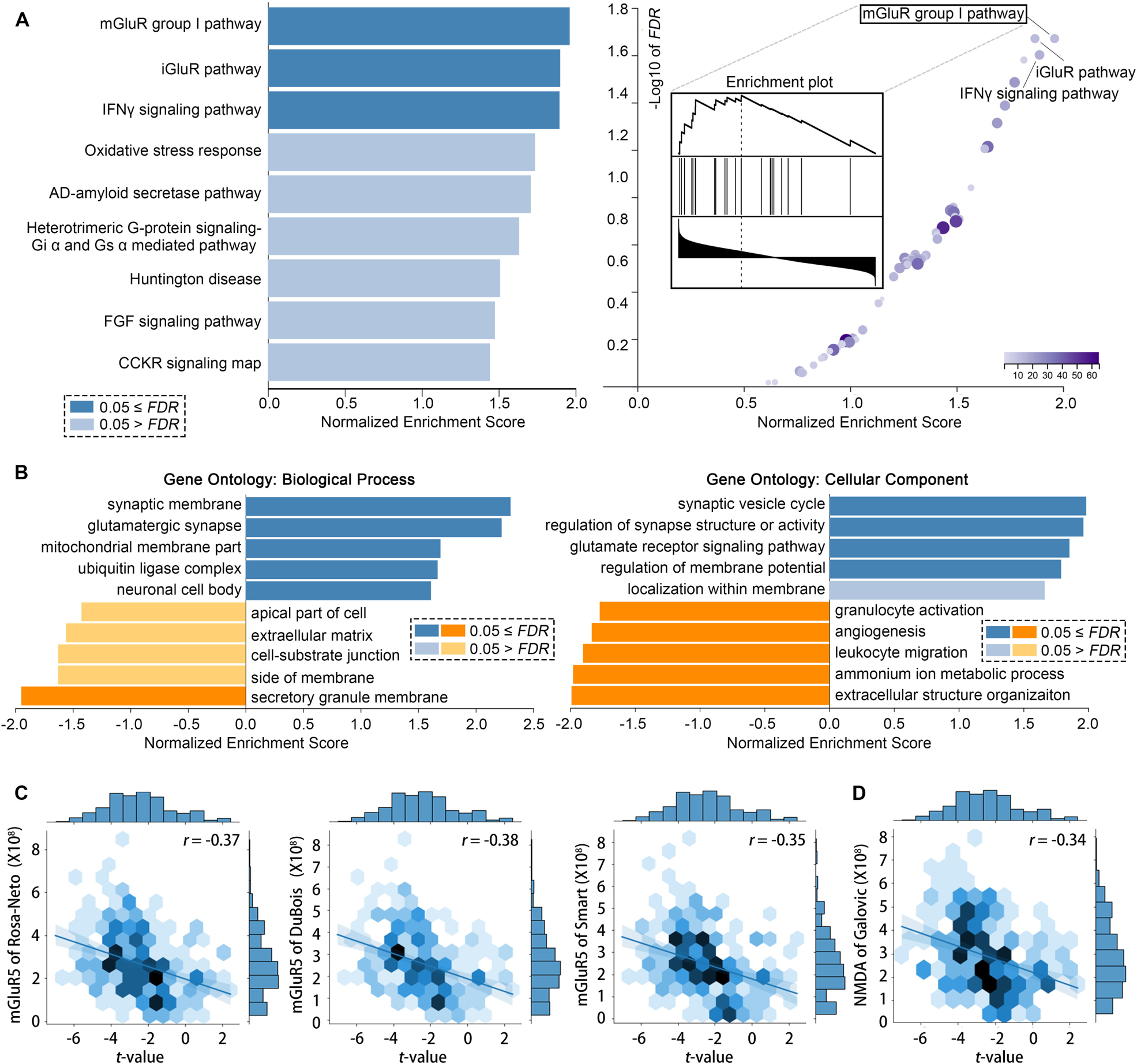
Linking structural narcolepsy topology to glutamatergic neuropathophysiology. (**A**) Significant results of Panther pathway terms in the functional enrichment analysis performed by WebGestalt. (**B**) Bar graph of GO enrichment analysis based on affinity propagation for narcolepsy-featured genes. (**C**) Further verification by glutamatergic PET/SPECT data.

As glutamatergic neuropathophysiology had the highest significance in functional analysis, we tried further verification by glutamatergic PET/SPECT data from unrelated healthy subjects. With 5,000 SA-preserving surrogate maps as null models, there were significant correlations between the global network degree and mGluR5 (*r* = −0.37, −0.38 and −0.35 respectively for Rosa-Neto, Dubois and Smart data, all *P* <□0.001; Fig. 3C) and NMDA (*r* = −0.34 for Galovic data, *P* <□0.001; Fig. 3D).

## 4. Discussion

In our present study, we explored the abnormal alteration in the overall structural network in people with narcolepsy, revealing a wide and significant wiring decline. It not only suggested a dramatic decrease in global efficiency of structural connectivity, but also showed a significant clinical correlation with shorter sleep latency, more frequently nocturnal awakening, and disrupted sleep macrostructure. It also showed that this structural alteration was closely linked to functional signature of glutamatergic neuropathophysiology, while additional PET/SPECT data verified that this structural alteration was significantly correlated with mGluR5 and NMDA. All these suggest that we need to pay more attention to the neuroplasticity features underlying the brain connectome adaptation in narcolepsy.

The human brain is a complex computational system which is widely connected by network of “highways” the information flow along. With the improvement of non-invasive imaging systems, much efforts toward investigating brain connectome focuses on the application of graph theoretical analysis, which facilitates explorations of the information propagation, integration, and segregation that characterize the topology of the network ^38^. It is widely reported in various neurological diseases, such as dementia, multiple sclerosis and stroke, that a bunch of architectural features are found sensitive to early disease diagnosis and symptom assessment, including efficiency ^39^, modularity ^40^, network hubs ^41^, and rich-club organization ^42^. In recent years, alterations in connectome have been found to be also sensitive to sleep disorders. For example, people with insomnia present a hyperconnectivity subnetwork related to right angular gyrus which contributes to increased reactivity and vulnerability ^43^; obstructive sleep apnea makes people more prone to connectivity dysfunction in the key nodes of the executive network and DMN ^44^. Most of studies on sleep disorder have focused on functional abnormalities, while structural abnormalities are reported infrequently. In our study, however, we found solid evidence that the structural networks in people with narcolepsy were remarkably damaged, so that the overall topological characteristics of the structural architecture were dramatically impaired. Although the topological metrics of people with narcolepsy presented little difference between subtypes, the descending structural properties of people with narcolepsy showed a close association with various symptoms (MLST sleep latency, awakening at night PSG and sleep macrostructure), suggesting an urge of studying the pathophysiological mechanism of the white matter damage in people with narcolepsy.

In addition to further mining structural biomarkers for narcolepsy, we also need to pay attention to the role that structural connectivity in shaping the brain activity dynamics of people with narcolepsy. Tremendous efforts in the early theoretical simulations relating structural and functional connectivity highlight that anatomical variability is one of the major factors that promote individualized static brain activity ^45,46^. And modelling functional connectivity has not only use of brain structural connectivity data of a healthy brain but also increasing need for that of brain disease ^47,48^. In the study of narcolepsy, adolescent with narcolepsy exhibit disrupted small-world network properties in functional activity as well as regional alterations in the caudate nucleus and posterior cingulate gyrus ^3^. The abnormal connectivities between the hypothalamus and brain regions involve in memory consolidation during sleep, such as the hippocampus, may be linked to the loss of hypocretin neurons in the dorsolateral hypothalamus ^49^. Further understanding the spatial properties, temporal dynamics, and spectral features of brain activity dynamics in narcolepsy asks for study deeper characteristics of the structural connectivity itself as well as structure-function relationship research.

One highlighted point during our connectome analysis is the anomalously enhanced subnetwork involving the thalamus, striatum, and CG that was closely associated with RBD. RBD is characterized by a loss of RWA together with prominent motor behavioral manifestations associated with dreaming during REM sleep 50. Narcolepsy seems to be the second most common cause of “secondary” RBD, accounting for about 10-15% of cases 51. However, whether it is an intrinsic feature or associated feature is a still unclear pathophysiology 52. Clinical studies have shown that narcolepsy RBD mainly affects children or young patients under the age of 50, with no difference in gender distribution, while idiopathic RBD or parkinsonism RBD is more common in middle-aged men 51. The incidence of RBD in narcolepsy type 1 ranged from 7% to 63% in different cohorts 52, while in some reports, the incidence of RBD in narcolepsy type 2 patients appeared to be much lower than that in narcolepsy type 1 (37% vs. 15%) 53. It is worth emphasizing the incidence of narcolepsy RBD can be easily overlooked, leading to an underestimation of the real prevalence 52. Overall, studies based on clinical interviews reported an approximate prevalence of 60-70%, which drops down to 2-50% in studies adopting video-PSG 52. And although these less violent RBD in narcolepsy may be an early symptom in childhood and preceded in severity and frequency with the aging process 54, there is currently no indication that they would develop symptoms or signs of neurodegenerative disorders as idiopathic RBD did 55. All these may suggest potential circuit mechanism differences between narcolepsy RBD and idiopathic RBD. In our present analysis, we found that the RBD-sensitive subnetwork in people with narcolepsy were mainly linked to CG, which was clearly different from the brainstem destruction (especially in sublaterodorsal tegmental nucleus) in idiopathic RBD 56,57. Therefore, it would be interesting to further characterize RBD-sensitive subnetwork in people with idiopathic RBD.

Further imaging genetics indicated that a glutamatergic neuropathophysiology mechanism may be underlying all above connectome remodeling in people with narcolepsy. It is well known that the hypocretin system regulates sleep/wake control via complex interactions with monoaminergic, cholinergic and GABA-ergic neuronal systems ^58^. The highly selective and severe loss of the hypocretinergic neurons promoted excessive daytime sleepiness, especially in the narcolepsy type 1 ^59^. Various studies have shown that the activity of hypocretin is closely related to glutamate: (1) During hypocretin releasing, it also attaches to glutaminergic receptor and induces positive feedback of following hypocretin release ^60,61^. (2) Hypocretinergic neurons release a lot of glutamate, which synergizes with the excitatory effects of hypocretin ^62,63^. When glutamate receptors are blocked, hypocretin does not exert its excitatory effect on motor neurons ^62,63^. (3) Hypocretinergic neurons also secrete neuronal activity-regulated pentraxin (NARP), a protein that promotes clustering of glutamatergic receptors at excitatory synapses and enhance postsynaptic responses to glutamate ^64,65^. Whether this glutamatergic neuropathophysiology is also involved in the development of narcolepsy subtype 2 has not been clearly reported. It’s worthy of our further exploration of their biological regulation within different narcolepsy subtypes.

One important limitation of this study is that most people with narcolepsy lacked hypocretin data. In recent years, more and more studies emphasize the importance of hypocretin in the pathogenesis of narcolepsy. In the text revision of ICSD-3 (ICSD-3-TR) released in 2023 (https://learn.aasm.org/Listing/a1341000002XmRvAAK), it is recommended that any potential person with narcolepsy should accept the hypocretin radioimmunoassay for diagnose and classification. However, there has been a considerable controversy over the standard of CSF hypocretin suitable for Chinese people with narcolepsy. Professor Fang Han pointed out in 2013 that the Chinese people’s CSF hypocretin was relatively lower than westerners ^66^. In his reports, the level of 138.0 ng/L may be the optimal cutoff for the diagnosis of people with narcolepsy type 1. Since that, his and other Chinese teams have kept trying to find other reliable alternative biomarkers for narcolepsy, such as DQB1*0602 ^67^. To deal with this problem, Professor Shuqin Zhan of Xuanwu Hospital initiated a multicenter study of clinical characterization and related biomarkers in narcolepsy in China since 2021 (ID: ChiCTR2100052316; public site: https://www.chictr.org.cn/showproj.html?proj=126978). We also join this project and hope it will provide a solid answer to the long-standing debate on the hypocretin criteria for Chinese people with narcolepsy. We will also conduct further structural study in the subsequent research based on the new optimal cutoff for CSF hypocretin for Chinese.

Several other methodological considerations should also be contemplated when interpreting the results of this study. Our findings are based on structural data determined using diffusion tractography. The limitation of this method includes the uncertainty of crossing fibers. There would be an improvement if high-angular resolution diffusion-weighted imaging or spherical deconvolution post-processing techniques are used to provide the best possible estimate of the underlying structural connectivity ^68^. Using transcriptome and PET/SPECT data from healthy human brains could be limited if the narcolepsy gene expression patterns are different from those of healthy brains.

## Data Availability

All data produced in the present study are available upon reasonable request to the authors.All code produced are available online at https://github.com/Jiaji-Lab.

## 5. Acknowledgments

This work has been supported by the Key R&D Program of Shaanxi Province (Grant No. 2022ZDLSF03-07).

