## Supplementary Material for "Disrupted topological properties of structural brain networks present a glutamatergic neuropathophysiology in people with narcolepsy"

### Supplementary Figure 1

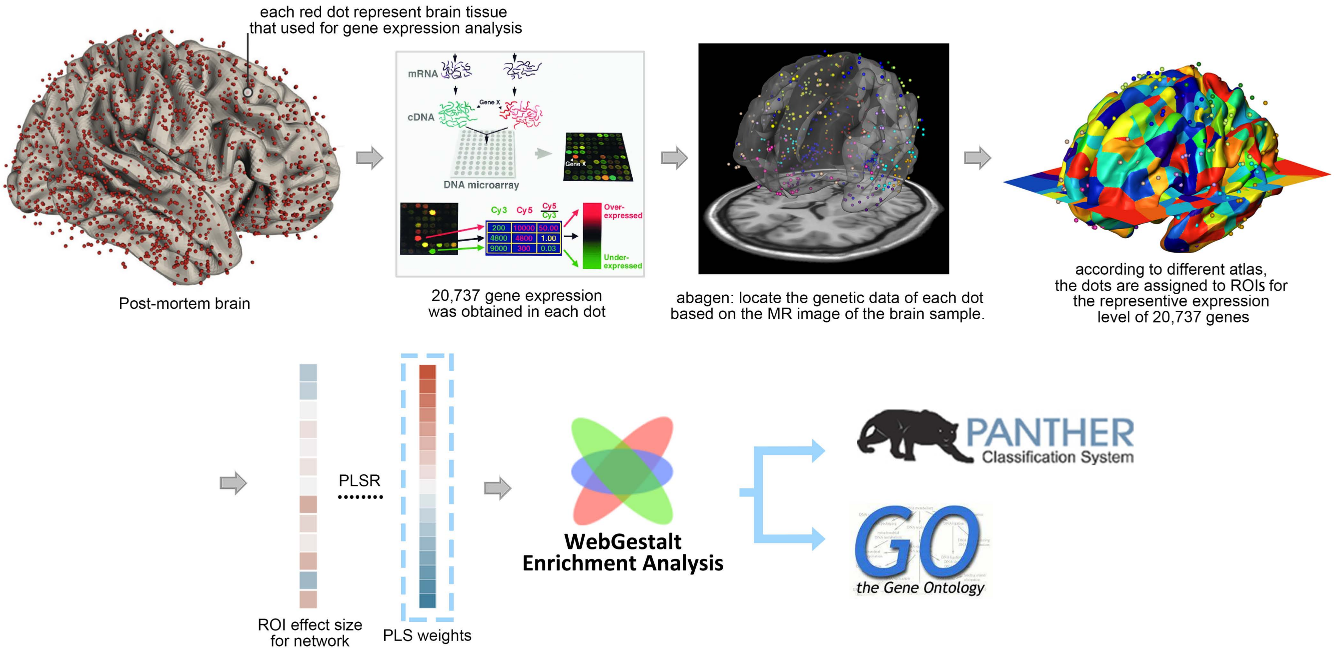

**Supplementary Figure 2**

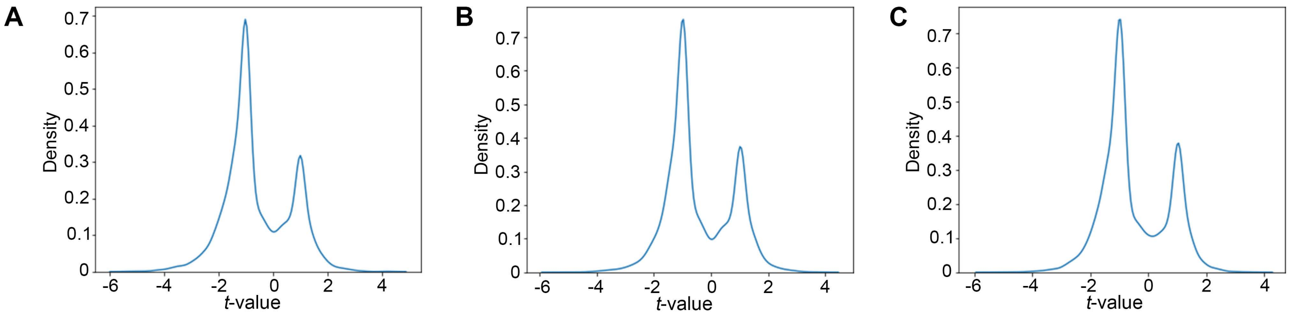


Figure Legend: (A) The t-values distribution between all NCs vs. their matched HCs; (B) The t-values distribution between all NC-1s vs. their matched HCs; (C) The t-values distribution between all NC-2s vs. their matched HCs.

**Supplementary Table 1 Top 20 Panther pathway terms for narcolepsy featured alterations**

| Lobe | Gyrus | Left and Right Hemisphere | Label ID.L | Label ID.R | Anatomical and modified Cyto-architectonic descriptions |
|---|---|---|---|---|---|
| **Frontal Lobe** | SFG, Superior Frontal Gyrus | SFG_L(R)_7_1 | 1 | 2 | *A8m, medial area 8* |
| | | SFG_L(R)_7_2 | 3 | 4 | *A8dl, dorsolateral area 8* |
| | | SFG_L(R)_7_3 | 5 | 6 | *A9l, lateral area 9* |
| | | SFG_L(R)_7_4 | 7 | 8 | *A6dl, dorsolateral area 6* |
| | | SFG_L(R)_7_5 | 9 | 10 | *A6m, medial area 6* |
| | | SFG_L(R)_7_6 | 11 | 12 | *A9m,medial area 9* |
| | | SFG_L(R)_7_7 | 13 | 14 | *A10m, medial area 10* |
| | MFG, Middle Frontal Gyrus | MFG_L(R)_7_1 | 15 | 16 | *A9/46d, dorsal area 9/46* |
| | | MFG_L(R)_7_2 | 17 | 18 | *IFJ, inferior frontal junction* |
| | | MFG_L(R)_7_3 | 19 | 20 | *A46, area 46* |
| | | MFG_L(R)_7_4 | 21 | 22 | *A9/46v, ventral area 9/46* |
| | | MFG_L(R)_7_5 | 23 | 24 | *A8vl, ventrolateral area 8* |
| | | MFG_L(R)_7_6 | 25 | 26 | *A6vl, ventrolateral area 6* |
| | | MFG_L(R)_7_7 | 27 | 28 | *A10l, lateral area10* |
| | IFG, Inferior Frontal Gyrus | IFG_L(R)_6_1 | 29 | 30 | *A44d,dorsal area 44* |
| | | IFG_L(R)_6_2 | 31 | 32 | *IFS, inferior frontal sulcus* |
| | | IFG_L(R)_6_3 | 33 | 34 | *A45c, caudal area 45* |
| | | IFG_L(R)_6_4 | 35 | 36 | *A45r, rostral area 45* |
| | | IFG_L(R)_6_5 | 37 | 38 | *A44op, opercular area 44* |
| | | IFG_L(R)_6_6 | 39 | 40 | *A44v, ventral area 44* |
| | OrG, Orbital Gyrus | OrG_L(R)_6_1 | 41 | 42 | *A14m, medial area 14* |
| | | OrG_L(R)_6_2 | 43 | 44 | *A12/47o, orbital area 12/47* |
| | | OrG_L(R)_6_3 | 45 | 46 | *A11l, lateral area 11* |
| | | OrG_L(R)_6_4 | 47 | 48 | *A11m, medial area 11* |
| | | OrG_L(R)_6_5 | 49 | 50 | *A13, area 13* |
| | | OrG_L(R)_6_6 | 51 | 52 | *A12/47l, lateral area 12/47* |
| | PrG, Precentral Gyrus | PrG_L(R)_6_1 | 53 | 54 | *A4hf, area 4(head and face region)* |
| | | PrG_L(R)_6_2 | 55 | 56 | *A6cdl, caudal dorsolateral area 6* |
| | | PrG_L(R)_6_3 | 57 | 58 | *A4ul, area 4(upper limb region)* |
| | | PrG_L(R)_6_4 | 59 | 60 | *A4t, area 4(trunk region)* |
| | | PrG_L(R)_6_5 | 61 | 62 | *A4tl, area 4(tongue and larynx region)* |

| | | | | | |
|---|---|---|---|---|---|
| | | PrG_L(R)_6_6 | 63 | 64 | *A6cvl, caudal ventrolateral area 6* |
| | PCL, Paracentral Lobule | PCL_L(R)_2_1 | 65 | 66 | *A1/2/3ll, area1/2/3 (lower limb region)* |
| | | PCL_L(R)_2_2 | 67 | 68 | *A4ll, area 4, (lower limb region)* |
| **Temporal Lobe** | STG, Superior Temporal Gyrus | STG_L(R)_6_1 | 69 | 70 | *A38m, medial area 38* |
| | | STG_L(R)_6_2 | 71 | 72 | *A41/42, area 41/42* |
| | | STG_L(R)_6_3 | 73 | 74 | *TE1.0 and TE1.2* |
| | | STG_L(R)_6_4 | 75 | 76 | *A22c, caudal area 22* |
| | | STG_L(R)_6_5 | 77 | 78 | *A38l, lateral area 38* |
| | | STG_L(R)_6_6 | 79 | 80 | *A22r, rostral area 22* |
| | MTG, Middle Temporal Gyrus | MTG_L(R)_4_1 | 81 | 82 | *A21c, caudal area 21* |
| | | MTG_L(R)_4_2 | 83 | 84 | *A21r, rostral area 21* |
| | | MTG_L(R)_4_3 | 85 | 86 | *A37dl, dorsolateral area37* |
| | | MTG_L(R)_4_4 | 87 | 88 | *aSTS, anterior superior temporal sulcus* |
| | ITG, Inferior Temporal Gyrus | ITG_L(R)_7_1 | 89 | 90 | *A20iv, intermediate ventral area 20* |
| | | ITG_L(R)_7_2 | 91 | 92 | *A37elv, extreme lateroventral area37* |
| | | ITG_L(R)_7_3 | 93 | 94 | *A20r, rostral area 20* |
| | | ITG_L(R)_7_4 | 95 | 96 | *A20il, intermediate lateral area 20* |
| | | ITG_L(R)_7_5 | 97 | 98 | *A37vl, ventrolateral area 37* |
| | | ITG_L(R)_7_6 | 99 | 100 | *A20cl, caudolateral of area 20* |
| | | ITG_L(R)_7_7 | 101 | 102 | *A20cv, caudoventral of area 20* |
| | FuG, Fusiform Gyrus | FuG_L(R)_3_1 | 103 | 104 | *A20rv, rostroventral area 20* |
| | | FuG_L(R)_3_2 | 105 | 106 | *A37mv, medioventral area37* |
| | | FuG_L(R)_3_3 | 107 | 108 | *A37lv, lateroventral area37* |
| | PhG, Parahippocampal Gyrus | PhG_L(R)_6_1 | 109 | 110 | *A35/36r, rostral area 35/36* |
| | | PhG_L(R)_6_2 | 111 | 112 | *A35/36c, caudal area 35/36* |
| | | PhG_L(R)_6_3 | 113 | 114 | *TL, area TL (lateral PPHC, posterior parahippocampal gyrus)* |
| | | PhG_L(R)_6_4 | 115 | 116 | *A28/34, area 28/34 (EC, entorhinal cortex)* |
| | | PhG_L(R)_6_5 | 117 | 118 | *TI, area TI(temporal* |

| | | | | | |
|---|---|---|---|---|---|
| | | | | | *agranular insular cortex)* |
| | | PhG_L(R)_6_6 | 119 | 120 | *TH, area TH (medial PPHC)* |
| | pSTS, posterior Superior Temporal Sulcus | pSTS_L(R)_2_1 | 121 | 122 | *rpSTS, rostroposterior superior temporal sulcus* |
| | | pSTS_L(R)_2_2 | 123 | 124 | *cpSTS, caudoposterior superior temporal sulcus* |
| **Parietal Lobe** | SPL, Superior Parietal Lobule | SPL_L(R)_5_1 | 125 | 126 | *A7r, rostral area 7* |
| | | SPL_L(R)_5_2 | 127 | 128 | *A7c, caudal area 7* |
| | | SPL_L(R)_5_3 | 129 | 130 | *A5l, lateral area 5* |
| | | SPL_L(R)_5_4 | 131 | 132 | *A7pc, postcentral area 7* |
| | | SPL_L(R)_5_5 | 133 | 134 | *A7ip, intraparietal area 7(hIP3)* |
| | IPL, Inferior Parietal Lobule | IPL_L(R)_6_1 | 135 | 136 | *A39c, caudal area 39(PGp)* |
| | | IPL_L(R)_6_2 | 137 | 138 | *A39rd, rostrodorsal area 39(Hip3)* |
| | | IPL_L(R)_6_3 | 139 | 140 | *A40rd, rostrodorsal area 40(PFt)* |
| | | IPL_L(R)_6_4 | 141 | 142 | *A40c, caudal area 40(PFm)* |
| | | IPL_L(R)_6_5 | 143 | 144 | *A39rv, rostroventral area 39(PGa)* |
| | | IPL_L(R)_6_6 | 145 | 146 | *A40rv, rostroventral area 40(PFop)* |
| | Pcun, Precuneus | PCun_L(R)_4_1 | 147 | 148 | *A7m, medial area 7(PEp)* |
| | | PCun_L(R)_4_2 | 149 | 150 | *A5m, medial area 5(PEm)* |
| | | PCun_L(R)_4_3 | 151 | 152 | *dmPOS, dorsomedial parietooccipital sulcus(PEr)* |
| | | PCun_L(R)_4_4 | 153 | 154 | *A31, area 31 (Lc1)* |
| | PoG, Postcentral Gyrus | PoG_L(R)_4_1 | 155 | 156 | *A1/2/3ulhf, area 1/2/3(upper limb, head and face region)* |
| | | PoG_L(R)_4_2 | 157 | 158 | *A1/2/3tonIa, area 1/2/3(tongue and larynx region)* |
| | | PoG_L(R)_4_3 | 159 | 160 | *A2, area 2* |
| | | PoG_L(R)_4_4 | 161 | 162 | *A1/2/3tru, area1/2/3(trunk region)* |
| **Insular Lobe** | INS, Insular Gyrus | INS_L(R)_6_1 | 163 | 164 | *G, hypergranular insula* |
| | | INS_L(R)_6_2 | 165 | 166 | *vIa, ventral agranular insula* |
| | | INS_L(R)_6_3 | 167 | 168 | *dIa, dorsal agranular insula* |
| | | INS_L(R)_6_4 | 169 | 170 | *vId/vIg, ventral dysgranular and granular insula* |

| | | | | | |
|---|---|---|---|---|---|
| | | INS_L(R)_6_5 | 171 | 172 | *dIg, dorsal granular insula* |
| | | INS_L(R)_6_6 | 173 | 174 | *dId, dorsal dysgranular insula* |
| **Limbic Lobe** | CG, Cingulate Gyrus | CG_L(R)_7_1 | 175 | 176 | *A23d, dorsal area 23* |
| | | CG_L(R)_7_2 | 177 | 178 | *A24rv, rostroventral area 24* |
| | | CG_L(R)_7_3 | 179 | 180 | *A32p, pregenual area 32* |
| | | CG_L(R)_7_4 | 181 | 182 | *A23v, ventral area 23* |
| | | CG_L(R)_7_5 | 183 | 184 | *A24cd, caudodorsal area 24* |
| | | CG_L(R)_7_6 | 185 | 186 | *A23c, caudal area 23* |
| | | CG_L(R)_7_7 | 187 | 188 | *A32sg, subgenual area 32* |
| **Occipital Lobe** | MVOcC, MedioVentral Occipital Cortex | MVOcC _L(R)_5_1 | 189 | 190 | *cLinG, caudal lingual gyrus* |
| | | MVOcC _L(R)_5_2 | 191 | 192 | *rCunG, rostral cuneus gyrus* |
| | | MVOcC _L(R)_5_3 | 193 | 194 | *cCunG, caudal cuneus gyrus* |
| | | MVOcC _L(R)_5_4 | 195 | 196 | *rLinG, rostral lingual gyrus* |
| | | MVOcC _L(R)_5_5 | 197 | 198 | *vmPOS,ventromedial parietooccipital sulcus* |
| | LOcC, lateral Occipital Cortex | LOcC_L(R)_4_1 | 199 | 200 | *mOccG, middle occipital gyrus* |
| | | LOcC _L(R)_4_2 | 201 | 202 | *V5/MT+, area V5/MT+* |
| | | LOcC _L(R)_4_3 | 203 | 204 | *OPC, occipital polar cortex* |
| | | LOcC_L(R)_4_4 | 205 | 206 | *iOccG, inferior occipital gyrus* |
| | | LOcC _L(R)_2_1 | 207 | 208 | *msOccG, medial superior occipital gyrus* |
| | | LOcC _L(R)_2_2 | 209 | 210 | *lsOccG, lateral superior occipital gyrus* |
| **Subcortical Nuclei** | Amyg, Amygdala | Amyg_L(R)_2_1 | 211 | 212 | *mAmyg, medial amygdala* |
| | | Amyg_L(R)_2_2 | 213 | 214 | *lAmyg, lateral amygdala* |
| | Hipp, Hippocampus | Hipp_L(R)_2_1 | 215 | 216 | *rHipp, rostral hippocampus* |
| | | Hipp_L(R)_2_2 | 217 | 218 | *cHipp, caudal hippocampus* |
| | BG, Basal Ganglia | BG_L(R)_6_1 | 219 | 220 | *vCa, ventral caudate* |
| | | BG_L(R)_6_2 | 221 | 222 | *GP, globus pallidus* |
| | | BG_L(R)_6_3 | 223 | 224 | *NAC, nucleus accumbens* |
| | | BG_L(R)_6_4 | 225 | 226 | *vmPu, ventromedial putamen* |
| | | BG_L(R)_6_5 | 227 | 228 | *dCa, dorsal caudate* |
| | | BG_L(R)_6_6 | 229 | 230 | *dlPu, dorsolateral putamen* |
| | Tha, Thalamus | Tha_L(R)_8_1 | 231 | 232 | *mPFtha, medial pre-frontal thalamus* |
| | | Tha_L(R)_8_2 | 233 | 234 | *mPMtha, pre-motor thalamus* |

| | | | | | |
|---|---|---|---|---|---|
| | | Tha_L(R)_8_3 | 235 | 236 | *Stha, sensory thalamus* |
| | | Tha_L(R)_8_4 | 237 | 238 | *rTtha, rostral temporal thalamus* |
| | | Tha_L(R)_8_5 | 239 | 240 | *PPtha, posterior parietal thalamus* |
| | | Tha_L(R)_8_6 | 241 | 242 | *Otha, occipital thalamus* |
| | | Tha_L(R)_8_7 | 243 | 244 | *cTtha, caudal temporal thalamus* |
| | | Tha_L(R)_8_8 | 245 | 246 | *lPFtha, lateral pre-frontal thalamus* |

**Supplementary Table 2 Nodal degree comparison**

| ROI | Brainnetome region | Subregion name | *t* value[a] (HC vs. NC) | *P*-adj[a] (HC vs. NC) | *t* value[b] (HC vs. NC-1) | *P*-adj[b] (HC vs. NC-1) | *t* value[c] (HC vs. NC-2) | *P*-adj[c] (HC vs. NC-2) |
|---|---|---|---|---|---|---|---|---|
| 1 | SFG_L_7_1 | A8m_L | 1.130 | 0.317 | 1.419 | 0.280 | 0.218 | 0.866 |
| 2 | SFG_R_7_1 | A8m_R | 1.127 | 0.317 | 1.516 | 0.244 | 0.345 | 0.787 |
| 3 | SFG_L_7_2 | A8dl_L | 2.260 | 0.055 | 2.538 | 0.060 | 0.769 | 0.562 |
| 4 | SFG_R_7_2 | A8dl_R | 4.883 | 0.000 | 3.045 | 0.039 | 3.933 | 0.030 |
| 5 | SFG_L_7_3 | A9l_L | 5.542 | 0.000 | 3.851 | 0.013 | 3.941 | 0.030 |
| 6 | SFG_R_7_3 | A9l_R | 3.454 | 0.006 | 2.027 | 0.125 | 3.130 | 0.048 |
| 7 | SFG_L_7_4 | A6dl_L | -0.664 | 0.565 | -0.997 | 0.451 | 0.092 | 0.932 |
| 8 | SFG_R_7_4 | A6dl_R | 0.301 | 0.805 | 0.060 | 0.961 | 0.357 | 0.781 |
| 9 | SFG_L_7_5 | A6m_L | -1.220 | 0.283 | -0.814 | 0.537 | -0.893 | 0.501 |
| 10 | SFG_R_7_5 | A6m_R | -0.162 | 0.891 | -0.629 | 0.642 | 0.452 | 0.727 |
| 11 | SFG_L_7_6 | A9m_L | 3.844 | 0.003 | 2.977 | 0.040 | 2.363 | 0.105 |
| 12 | SFG_R_7_6 | A9m_R | 2.992 | 0.014 | 2.168 | 0.100 | 2.093 | 0.131 |
| 13 | SFG_L_7_7 | A10m_L | 3.744 | 0.003 | 2.770 | 0.049 | 2.497 | 0.091 |
| 14 | SFG_R_7_7 | A10m_R | 3.838 | 0.003 | 2.249 | 0.087 | 3.277 | 0.046 |
| 15 | MFG_L_7_1 | A9/46d_L | 5.206 | 0.000 | 4.228 | 0.011 | 3.217 | 0.046 |
| 16 | MFG_R_7_1 | A9/46d_R | 3.938 | 0.002 | 3.488 | 0.021 | 1.957 | 0.156 |
| 17 | MFG_L_7_2 | IFJ_L | 2.395 | 0.044 | 2.015 | 0.127 | 1.260 | 0.340 |
| 18 | MFG_R_7_2 | IFJ_R | 3.950 | 0.002 | 3.461 | 0.021 | 2.140 | 0.127 |
| 19 | MFG_L_7_3 | A46_L | 3.032 | 0.013 | 4.098 | 0.011 | 0.743 | 0.575 |
| 20 | MFG_R_7_3 | A46_R | 5.997 | 0.000 | 4.059 | 0.011 | 5.029 | 0.024 |
| 21 | MFG_L_7_4 | A9/46v_L | 4.526 | 0.001 | 4.083 | 0.011 | 2.510 | 0.091 |
| 22 | MFG_R_7_4 | A9/46v_R | 4.876 | 0.000 | 3.081 | 0.037 | 3.920 | 0.030 |
| 23 | MFG_L_7_5 | A8vl_L | 5.613 | 0.000 | 4.884 | 0.011 | 3.212 | 0.046 |
| 24 | MFG_R_7_5 | A8vl_R | 6.256 | 0.000 | 4.282 | 0.011 | 4.536 | 0.028 |
| 25 | MFG_L_7_6 | A6vl_L | 3.414 | 0.006 | 1.803 | 0.173 | 3.076 | 0.049 |
| 26 | MFG_R_7_6 | A6vl_R | 3.885 | 0.003 | 3.364 | 0.025 | 2.176 | 0.123 |
| 27 | MFG_L_7_7 | A10l_L | 3.530 | 0.005 | 2.652 | 0.057 | 2.336 | 0.108 |
| 28 | MFG_R_7_7 | A10l_R | 4.233 | 0.001 | 2.607 | 0.059 | 3.447 | 0.042 |
| 29 | IFG_L_6_1 | A44d_L | 2.674 | 0.026 | 2.595 | 0.059 | 1.373 | 0.294 |
| 30 | IFG_R_6_1 | A44d_R | 2.075 | 0.073 | 1.826 | 0.168 | 1.130 | 0.382 |
| 31 | IFG_L_6_2 | IFS_L | 5.479 | 0.000 | 3.656 | 0.015 | 4.137 | 0.028 |
| 32 | IFG_R_6_2 | IFS_R | 3.129 | 0.011 | 2.121 | 0.109 | 2.256 | 0.116 |
| 33 | IFG_L_6_3 | A45c_L | 4.676 | 0.001 | 4.184 | 0.011 | 2.426 | 0.101 |
| 34 | IFG_R_6_3 | A45c_R | 4.399 | 0.001 | 3.999 | 0.012 | 2.128 | 0.127 |
| 35 | IFG_L_6_4 | A45r_L | 2.748 | 0.023 | 3.117 | 0.035 | 1.177 | 0.370 |
| 36 | IFG_R_6_4 | A45r_R | 4.910 | 0.000 | 2.893 | 0.042 | 4.354 | 0.028 |
| 37 | IFG_L_6_5 | A44op_L | 4.515 | 0.001 | 4.560 | 0.011 | 2.411 | 0.102 |
| 38 | IFG_R_6_5 | A44op_R | 5.056 | 0.000 | 3.816 | 0.013 | 3.190 | 0.047 |
| 39 | IFG_L_6_6 | A44v_L | 3.100 | 0.011 | 2.759 | 0.049 | 1.695 | 0.216 |
| 40 | IFG_R_6_6 | A44v_R | 4.867 | 0.000 | 3.266 | 0.028 | 3.572 | 0.038 |
| 41 | OrG_L_6_1 | A14m_L | 3.858 | 0.003 | 2.571 | 0.059 | 3.020 | 0.052 |
| 42 | OrG_R_6_1 | A14m_R | 0.034 | 0.981 | -0.334 | 0.796 | 0.600 | 0.649 |
| 43 | OrG_L_6_2 | A12/47o_L | 3.297 | 0.007 | 2.514 | 0.061 | 2.237 | 0.117 |
| 44 | OrG_R_6_2 | A12/47o_R | 5.543 | 0.000 | 4.651 | 0.011 | 3.244 | 0.046 |
| 45 | OrG_L_6_3 | A11l_L | 2.310 | 0.051 | 0.567 | 0.685 | 2.713 | 0.072 |

| | | | | | | | | |
|---|---|---|---|---|---|---|---|---|
| 46 | OrG_R_6_3 | A11l_R | 3.691 | 0.004 | 1.857 | 0.161 | 3.551 | 0.038 |
| 47 | OrG_L_6_4 | A11m_L | 4.099 | 0.002 | 2.506 | 0.061 | 3.315 | 0.045 |
| 48 | OrG_R_6_4 | A11m_R | 3.724 | 0.003 | 3.516 | 0.020 | 1.625 | 0.227 |
| 49 | OrG_L_6_5 | A13_L | 3.823 | 0.003 | 0.993 | 0.451 | 8.278 | 0.001 |
| 50 | OrG_R_6_5 | A13_R | 3.588 | 0.004 | 3.264 | 0.028 | 1.716 | 0.210 |
| 51 | OrG_L_6_6 | A12/47l_L | 4.798 | 0.001 | 3.004 | 0.040 | 4.395 | 0.028 |
| 52 | OrG_R_6_6 | A12/47l_R | 3.879 | 0.003 | 3.253 | 0.028 | 2.245 | 0.116 |
| 53 | PrG_L_6_1 | A4hf_L | 2.147 | 0.065 | 1.067 | 0.420 | 1.960 | 0.156 |
| 54 | PrG_R_6_1 | A4hf_R | 3.632 | 0.004 | 2.540 | 0.060 | 2.727 | 0.072 |
| 55 | PrG_L_6_2 | A6cdl_L | 0.971 | 0.395 | -0.548 | 0.690 | 1.574 | 0.233 |
| 56 | PrG_R_6_2 | A6cdl_R | -0.697 | 0.544 | 0.354 | 0.792 | -1.169 | 0.370 |
| 57 | PrG_L_6_3 | A4ul_L | -1.341 | 0.238 | -2.586 | 0.059 | 0.403 | 0.755 |
| 58 | PrG_R_6_3 | A4ul_R | -0.013 | 0.990 | 0.117 | 0.924 | -0.116 | 0.917 |
| 59 | PrG_L_6_4 | A4t_L | -2.428 | 0.042 | -2.640 | 0.057 | -1.234 | 0.350 |
| 60 | PrG_R_6_4 | A4t_R | -1.947 | 0.093 | -2.582 | 0.059 | -0.481 | 0.715 |
| 61 | PrG_L_6_5 | A4tl_L | 3.328 | 0.007 | 2.725 | 0.051 | 2.086 | 0.131 |
| 62 | PrG_R_6_5 | A4tl_R | 3.325 | 0.007 | 4.284 | 0.011 | 1.374 | 0.294 |
| 63 | PrG_L_6_6 | A6cvl_L | 3.901 | 0.003 | 2.933 | 0.041 | 2.507 | 0.091 |
| 64 | PrG_R_6_6 | A6cvl_R | 4.118 | 0.002 | 1.678 | 0.200 | 4.785 | 0.027 |
| 65 | PCL_L_2_1 | A1/2/3ll_L | 1.574 | 0.173 | 0.382 | 0.783 | 1.766 | 0.200 |
| 66 | PCL_R_2_1 | A1/2/3ll_R | -1.798 | 0.120 | -1.303 | 0.320 | -1.380 | 0.294 |
| 67 | PCL_L_2_2 | A4ll_L | 1.906 | 0.100 | 1.634 | 0.208 | 1.131 | 0.382 |
| 68 | PCL_R_2_2 | A4ll_R | 2.261 | 0.055 | 0.884 | 0.502 | 3.368 | 0.043 |
| 69 | STG_L_6_1 | A38m_L | 5.128 | 0.000 | 4.377 | 0.011 | 3.167 | 0.048 |
| 70 | STG_R_6_1 | A38m_R | 6.997 | 0.000 | 5.597 | 0.010 | 4.102 | 0.028 |
| 71 | STG_L_6_2 | A41/42_L | 5.673 | 0.000 | 4.134 | 0.011 | 3.910 | 0.030 |
| 72 | STG_R_6_2 | A41/42_R | 3.157 | 0.010 | 2.252 | 0.087 | 2.154 | 0.125 |
| 73 | STG_L_6_3 | TE1.0/TE1.2_L | 1.764 | 0.126 | 0.587 | 0.673 | 1.990 | 0.149 |
| 74 | STG_R_6_3 | TE1.0/TE1.2_R | 3.379 | 0.006 | 2.765 | 0.049 | 2.106 | 0.130 |
| 75 | STG_L_6_4 | A22c_L | 2.728 | 0.024 | 1.166 | 0.380 | 3.795 | 0.030 |
| 76 | STG_R_6_4 | A22c_R | 4.305 | 0.001 | 4.938 | 0.011 | 2.294 | 0.113 |
| 77 | STG_L_6_5 | A38l_L | 1.171 | 0.303 | 0.951 | 0.468 | 0.655 | 0.616 |
| 78 | STG_R_6_5 | A38l_R | 1.965 | 0.090 | 0.543 | 0.690 | 3.822 | 0.030 |
| 79 | STG_L_6_6 | A22r_L | 2.975 | 0.014 | 0.956 | 0.467 | 3.744 | 0.031 |
| 80 | STG_R_6_6 | A22r_R | 3.132 | 0.011 | 1.683 | 0.200 | 3.143 | 0.048 |
| 81 | MTG_L_4_1 | A21c_L | 3.944 | 0.002 | 3.290 | 0.028 | 2.253 | 0.116 |
| 82 | MTG_R_4_1 | A21c_R | 4.299 | 0.001 | 2.950 | 0.041 | 3.118 | 0.048 |
| 83 | MTG_L_4_2 | A21r_L | 3.488 | 0.005 | 2.803 | 0.048 | 2.176 | 0.123 |
| 84 | MTG_R_4_2 | A21r_R | 5.430 | 0.000 | 3.999 | 0.012 | 3.553 | 0.038 |
| 85 | MTG_L_4_3 | A37dl_L | 2.627 | 0.028 | 2.474 | 0.064 | 1.037 | 0.428 |
| 86 | MTG_R_4_3 | A37dl_R | 2.230 | 0.058 | 1.798 | 0.173 | 1.483 | 0.262 |
| 87 | MTG_L_4_4 | aSTS_L | 2.255 | 0.055 | 1.081 | 0.416 | 2.636 | 0.081 |
| 88 | MTG_R_4_4 | aSTS_R | 4.755 | 0.001 | 2.768 | 0.049 | 4.136 | 0.028 |
| 89 | ITG_L_7_1 | A20iv_L | 3.798 | 0.003 | 3.009 | 0.040 | 2.524 | 0.091 |
| 90 | ITG_R_7_1 | A20iv_R | 2.745 | 0.023 | 2.538 | 0.060 | 1.481 | 0.262 |
| 91 | ITG_L_7_2 | A37elv_L | 2.722 | 0.024 | 1.373 | 0.295 | 2.453 | 0.097 |
| 92 | ITG_R_7_2 | A37elv_R | 1.438 | 0.208 | 0.117 | 0.924 | 2.007 | 0.146 |
| 93 | ITG_L_7_3 | A20r_L | 2.212 | 0.059 | 1.210 | 0.362 | 2.209 | 0.119 |
| 94 | ITG_R_7_3 | A20r_R | 2.329 | 0.050 | 2.314 | 0.079 | 0.793 | 0.550 |

| | | | | | | | | |
|---|---|---|---|---|---|---|---|---|
| 95 | ITG_L_7_4 | A20il_L | 1.143 | 0.311 | 0.752 | 0.572 | 0.839 | 0.527 |
| 96 | ITG_R_7_4 | A20il_R | 5.626 | 0.000 | 4.231 | 0.011 | 3.831 | 0.030 |
| 97 | ITG_L_7_5 | A37vl_L | 2.491 | 0.037 | 4.128 | 0.011 | 0.153 | 0.900 |
| 98 | ITG_R_7_5 | A37vl_R | 1.372 | 0.231 | 0.544 | 0.690 | 1.279 | 0.333 |
| 99 | ITG_L_7_6 | A20cl_L | 2.690 | 0.025 | 1.705 | 0.194 | 2.111 | 0.130 |
| 100 | ITG_R_7_6 | A20cl_R | 3.867 | 0.003 | 3.942 | 0.013 | 2.007 | 0.146 |
| 101 | ITG_L_7_7 | A20cv_L | 2.290 | 0.053 | 0.524 | 0.699 | 3.113 | 0.048 |
| 102 | ITG_R_7_7 | A20cv_R | 2.616 | 0.029 | 3.396 | 0.024 | 0.134 | 0.911 |
| 103 | FuG_L_3_1 | A20rv_L | 3.144 | 0.010 | 2.659 | 0.057 | 1.685 | 0.217 |
| 104 | FuG_R_3_1 | A20rv_R | 3.947 | 0.002 | 2.543 | 0.060 | 3.041 | 0.051 |
| 105 | FuG_L_3_2 | A37mv_L | 4.013 | 0.002 | 2.617 | 0.058 | 3.024 | 0.052 |
| 106 | FuG_R_3_2 | A37mv_R | 2.139 | 0.065 | 2.035 | 0.125 | 1.023 | 0.435 |
| 107 | FuG_L_3_3 | A37lv_L | 4.206 | 0.002 | 4.431 | 0.011 | 1.618 | 0.227 |
| 108 | FuG_R_3_3 | A37lv_R | 3.268 | 0.008 | 3.241 | 0.028 | 1.606 | 0.227 |
| 109 | PhG_L_6_1 | A35/36r_L | 1.872 | 0.105 | 1.212 | 0.362 | 1.434 | 0.279 |
| 110 | PhG_R_6_1 | A35/36r_R | 3.438 | 0.006 | 2.345 | 0.075 | 2.522 | 0.091 |
| 111 | PhG_L_6_2 | A35/36c_L | 1.371 | 0.231 | 0.340 | 0.796 | 2.085 | 0.131 |
| 112 | PhG_R_6_2 | A35/36c_R | 2.264 | 0.055 | 1.657 | 0.201 | 2.501 | 0.091 |
| 113 | PhG_L_6_3 | TL_L | 1.150 | 0.310 | 1.602 | 0.219 | -0.152 | 0.900 |
| 114 | PhG_R_6_3 | TL_R | 1.173 | 0.303 | 0.333 | 0.796 | 1.155 | 0.375 |
| 115 | PhG_L_6_4 | A28/34_L | 2.589 | 0.030 | 2.311 | 0.079 | 1.226 | 0.351 |
| 116 | PhG_R_6_4 | A28/34_R | 1.502 | 0.195 | 0.838 | 0.525 | 1.606 | 0.227 |
| 117 | PhG_L_6_5 | TI_L | 1.103 | 0.327 | 0.687 | 0.608 | 0.883 | 0.505 |
| 118 | PhG_R_6_5 | TI_R | 1.356 | 0.236 | 0.424 | 0.757 | 1.842 | 0.184 |
| 119 | PhG_L_6_6 | TH_L | 2.200 | 0.059 | 1.369 | 0.295 | 2.379 | 0.104 |
| 120 | PhG_R_6_6 | TH_R | 2.173 | 0.062 | 1.108 | 0.402 | 2.598 | 0.086 |
| 121 | pSTS_L_2_1 | rpSTS_L | 2.259 | 0.055 | 0.991 | 0.451 | 2.338 | 0.108 |
| 122 | pSTS_R_2_1 | rpSTS_R | 0.757 | 0.511 | 0.542 | 0.690 | 0.513 | 0.701 |
| 123 | pSTS_L_2_2 | cpSTS_L | 2.870 | 0.018 | 1.075 | 0.418 | 3.258 | 0.046 |
| 124 | pSTS_R_2_2 | cpSTS_R | 3.666 | 0.004 | 3.716 | 0.014 | 2.072 | 0.133 |
| 125 | SPL_L_5_1 | A7r_L | 0.501 | 0.666 | 0.240 | 0.851 | 0.429 | 0.741 |
| 126 | SPL_R_5_1 | A7r_R | 2.976 | 0.014 | 2.708 | 0.052 | 1.587 | 0.229 |
| 127 | SPL_L_5_2 | A7c_L | 4.320 | 0.001 | 3.756 | 0.014 | 2.272 | 0.114 |
| 128 | SPL_R_5_2 | A7c_R | 1.225 | 0.281 | 0.635 | 0.642 | 1.301 | 0.325 |
| 129 | SPL_L_5_3 | A5l_L | -0.766 | 0.508 | 0.417 | 0.759 | -1.503 | 0.257 |
| 130 | SPL_R_5_3 | A5l_R | 2.712 | 0.024 | 1.721 | 0.191 | 2.839 | 0.063 |
| 131 | SPL_L_5_4 | A7pc_L | 2.079 | 0.072 | 1.054 | 0.426 | 1.899 | 0.169 |
| 132 | SPL_R_5_4 | A7pc_R | -0.567 | 0.623 | 0.248 | 0.848 | -0.984 | 0.451 |
| 133 | SPL_L_5_5 | A7ip_L | 1.462 | 0.204 | 0.117 | 0.924 | 1.682 | 0.217 |
| 134 | SPL_R_5_5 | A7ip_R | 1.686 | 0.146 | 3.031 | 0.039 | 0.653 | 0.616 |
| 135 | IPL_L_6_1 | A39c_L | 3.162 | 0.010 | 2.885 | 0.042 | 1.425 | 0.282 |
| 136 | IPL_R_6_1 | A39c_R | 3.511 | 0.005 | 4.358 | 0.011 | 1.231 | 0.350 |
| 137 | IPL_L_6_2 | A39rd_L | 2.714 | 0.024 | 2.429 | 0.066 | 1.371 | 0.294 |
| 138 | IPL_R_6_2 | A39rd_R | 2.786 | 0.022 | 2.075 | 0.118 | 1.787 | 0.196 |
| 139 | IPL_L_6_3 | A40rd_L | 5.159 | 0.000 | 3.854 | 0.013 | 3.431 | 0.042 |
| 140 | IPL_R_6_3 | A40rd_R | 2.186 | 0.061 | 1.864 | 0.160 | 1.170 | 0.370 |
| 141 | IPL_L_6_4 | A40c_L | 3.106 | 0.011 | 2.362 | 0.073 | 2.206 | 0.119 |
| 142 | IPL_R_6_4 | A40c_R | 3.107 | 0.011 | 1.753 | 0.183 | 2.710 | 0.072 |
| 143 | IPL_L_6_5 | A39rv_L | 3.441 | 0.006 | 3.720 | 0.014 | 1.640 | 0.226 |

| | | | | | | | | |
|---|---|---|---|---|---|---|---|---|
| 144 | IPL_R_6_5 | A39rv_R | 4.440 | 0.001 | 3.280 | 0.028 | 3.106 | 0.048 |
| 145 | IPL_L_6_6 | A40rv_L | 4.166 | 0.002 | 2.545 | 0.060 | 3.380 | 0.043 |
| 146 | IPL_R_6_6 | A40rv_R | 5.647 | 0.000 | 4.856 | 0.011 | 3.006 | 0.052 |
| 147 | PCun_L_4_1 | A7m_L | 4.190 | 0.002 | 2.898 | 0.042 | 2.952 | 0.055 |
| 148 | PCun_R_4_1 | A7m_R | 2.122 | 0.067 | 1.477 | 0.257 | 1.649 | 0.226 |
| 149 | PCun_L_4_2 | A5m_L | 2.209 | 0.059 | 2.506 | 0.061 | 0.588 | 0.654 |
| 150 | PCun_R_4_2 | A5m_R | 2.373 | 0.046 | 2.423 | 0.066 | 0.675 | 0.607 |
| 151 | PCun_L_4_3 | dmPOS_L | 2.220 | 0.058 | 0.198 | 0.875 | 4.168 | 0.028 |
| 152 | PCun_R_4_3 | dmPOS_R | 2.880 | 0.018 | 1.574 | 0.227 | 2.528 | 0.091 |
| 153 | PCun_L_4_4 | A31_L | -0.876 | 0.446 | -1.212 | 0.362 | 0.125 | 0.914 |
| 154 | PCun_R_4_4 | A31_R | 1.546 | 0.182 | 0.701 | 0.600 | 1.762 | 0.200 |
| 155 | PoG_L_4_1 | A1/2/3ulhf_L | 4.336 | 0.001 | 3.141 | 0.034 | 2.882 | 0.060 |
| 156 | PoG_R_4_1 | A1/2/3ulhf_R | 2.646 | 0.027 | 1.320 | 0.313 | 2.380 | 0.104 |
| 157 | PoG_L_4_2 | A1/2/3tonIa_L | 3.818 | 0.003 | 2.920 | 0.041 | 2.364 | 0.105 |
| 158 | PoG_R_4_2 | A1/2/3tonIa_R | 1.707 | 0.140 | 1.185 | 0.372 | 1.189 | 0.368 |
| 159 | PoG_L_4_3 | A2_L | 2.611 | 0.029 | 1.363 | 0.295 | 2.287 | 0.113 |
| 160 | PoG_R_4_3 | A2_R | 3.330 | 0.007 | 2.923 | 0.041 | 1.645 | 0.226 |
| 161 | PoG_L_4_4 | A1/2/3tru_L | 1.769 | 0.126 | 1.486 | 0.256 | 0.972 | 0.456 |
| 162 | PoG_R_4_4 | A1/2/3tru_R | -0.323 | 0.791 | -0.021 | 0.984 | -0.406 | 0.755 |
| 163 | INS_L_6_1 | G_L | 2.242 | 0.056 | 1.363 | 0.295 | 1.760 | 0.200 |
| 164 | INS_R_6_1 | G_R | 3.365 | 0.007 | 3.881 | 0.013 | 0.617 | 0.640 |
| 165 | INS_L_6_2 | vIa_L | 2.004 | 0.084 | 1.131 | 0.391 | 1.650 | 0.226 |
| 166 | INS_R_6_2 | vIa_R | 3.868 | 0.003 | 2.863 | 0.043 | 2.561 | 0.090 |
| 167 | INS_L_6_3 | dIa_L | 3.551 | 0.005 | 2.972 | 0.040 | 2.389 | 0.104 |
| 168 | INS_R_6_3 | dIa_R | 3.197 | 0.009 | 2.757 | 0.049 | 1.812 | 0.192 |
| 169 | INS_L_6_4 | vId/vIg_L | 1.376 | 0.231 | 0.904 | 0.492 | 1.143 | 0.379 |
| 170 | INS_R_6_4 | vId/vIg_R | 2.348 | 0.048 | 1.866 | 0.160 | 1.373 | 0.294 |
| 171 | INS_L_6_5 | dIg_L | -0.403 | 0.735 | -0.329 | 0.796 | -0.222 | 0.866 |
| 172 | INS_R_6_5 | dIg_R | 1.246 | 0.274 | 0.330 | 0.796 | 1.588 | 0.229 |
| 173 | INS_L_6_6 | dId_L | 1.891 | 0.102 | 1.946 | 0.142 | 0.712 | 0.588 |
| 174 | INS_R_6_6 | dId_R | 1.920 | 0.098 | 1.569 | 0.228 | 1.065 | 0.414 |
| 175 | CG_L_7_1 | A23d_L | 0.972 | 0.395 | 0.772 | 0.563 | 0.585 | 0.654 |
| 176 | CG_R_7_1 | A23d_R | 2.151 | 0.065 | 1.540 | 0.236 | 1.449 | 0.275 |
| 177 | CG_L_7_2 | A24rv_L | 1.574 | 0.173 | 1.134 | 0.391 | 1.616 | 0.227 |
| 178 | CG_R_7_2 | A24rv_R | -0.736 | 0.520 | -0.518 | 0.699 | -0.792 | 0.550 |
| 179 | CG_L_7_3 | A32p_L | 2.293 | 0.053 | 1.039 | 0.432 | 2.756 | 0.069 |
| 180 | CG_R_7_3 | A32p_R | 0.221 | 0.851 | 0.928 | 0.480 | -0.470 | 0.720 |
| 181 | CG_L_7_4 | A23v_L | 0.048 | 0.974 | -0.358 | 0.792 | 0.309 | 0.806 |
| 182 | CG_R_7_4 | A23v_R | 4.237 | 0.001 | 2.932 | 0.041 | 3.353 | 0.043 |
| 183 | CG_L_7_5 | A24cd_L | 2.203 | 0.059 | 2.645 | 0.057 | 0.845 | 0.526 |
| 184 | CG_R_7_5 | A24cd_R | 2.420 | 0.042 | 2.441 | 0.065 | 0.750 | 0.573 |
| 185 | CG_L_7_6 | A23c_L | -2.130 | 0.066 | -1.889 | 0.155 | -1.186 | 0.368 |
| 186 | CG_R_7_6 | A23c_R | -1.883 | 0.103 | -0.478 | 0.721 | -2.286 | 0.113 |
| 187 | CG_L_7_7 | A32sg_L | -0.271 | 0.822 | -0.823 | 0.534 | 0.791 | 0.550 |
| 188 | CG_R_7_7 | A32sg_R | 2.512 | 0.035 | 1.366 | 0.295 | 2.310 | 0.112 |
| 189 | MVOcC_L_5_1 | cLinG_L | 2.697 | 0.025 | 1.889 | 0.155 | 1.862 | 0.179 |
| 190 | MVOcC_R_5_1 | cLinG_R | 3.691 | 0.004 | 2.423 | 0.066 | 2.769 | 0.069 |
| 191 | MVOcC_L_5_2 | rCunG_L | 3.460 | 0.006 | 1.795 | 0.173 | 3.570 | 0.038 |
| 192 | MVOcC_R_5_2 | rCunG_R | 2.697 | 0.025 | 1.656 | 0.201 | 2.162 | 0.124 |

| | | | | | | | | |
|---|---|---|---|---|---|---|---|---|
| 193 | MVOcC_L_5_3 | cCunG_L | 1.456 | 0.204 | 0.377 | 0.784 | 2.515 | 0.091 |
| 194 | MVOcC_R_5_3 | cCunG_R | 3.614 | 0.004 | 2.584 | 0.059 | 2.810 | 0.064 |
| 195 | MVOcC_L_5_4 | rLinG_L | 4.054 | 0.002 | 1.952 | 0.142 | 4.135 | 0.028 |
| 196 | MVOcC_R_5_4 | rLinG_R | 2.416 | 0.042 | 2.463 | 0.064 | 1.627 | 0.227 |
| 197 | MVOcC_L_5_5 | vmPOS_L | 3.730 | 0.003 | 0.996 | 0.451 | 5.544 | 0.016 |
| 198 | MVOcC_R_5_5 | vmPOS_R | 2.539 | 0.033 | 0.993 | 0.451 | 2.955 | 0.055 |
| 199 | LOcC_L_4_1 | mOccG_L | 2.751 | 0.023 | 2.459 | 0.064 | 1.408 | 0.286 |
| 200 | LOcC_R_4_1 | mOccG_R | 3.277 | 0.008 | 2.037 | 0.125 | 2.937 | 0.055 |
| 201 | LOcC_L_4_2 | V5/MT+_L | 2.462 | 0.039 | 2.633 | 0.057 | 0.875 | 0.509 |
| 202 | LOcC_R_4_2 | V5/MT+_R | 3.030 | 0.013 | 3.909 | 0.013 | 1.603 | 0.227 |
| 203 | LOcC_L_4_3 | OPC_L | 2.457 | 0.039 | 1.704 | 0.194 | 1.755 | 0.200 |
| 204 | LOcC_R_4_3 | OPC_R | 3.566 | 0.005 | 2.801 | 0.048 | 2.174 | 0.123 |
| 205 | LOcC_L_4_4 | iOccG_L | 3.101 | 0.011 | 2.472 | 0.064 | 1.804 | 0.193 |
| 206 | LOcC_R_4_4 | iOccG_R | 3.549 | 0.005 | 2.196 | 0.096 | 2.835 | 0.063 |
| 207 | LOcC_L_2_1 | msOccG_L | 4.268 | 0.001 | 2.995 | 0.040 | 3.255 | 0.046 |
| 208 | LOcC_R_2_1 | msOccG_R | 3.983 | 0.002 | 3.827 | 0.013 | 1.721 | 0.210 |
| 209 | LOcC_L_2_2 | lsOccG_L | 4.443 | 0.001 | 4.121 | 0.011 | 2.223 | 0.118 |
| 210 | LOcC_R_2_2 | lsOccG_R | 3.866 | 0.003 | 2.534 | 0.060 | 2.892 | 0.059 |
| 211 | Amyg_L_2_1 | mAmyg_L | 1.778 | 0.124 | 0.459 | 0.732 | 2.130 | 0.127 |
| 212 | Amyg_R_2_1 | mAmyg_R | 1.480 | 0.202 | 0.484 | 0.720 | 3.351 | 0.043 |
| 213 | Amyg_L_2_2 | lAmyg_L | 0.262 | 0.825 | -1.140 | 0.390 | 1.361 | 0.299 |
| 214 | Amyg_R_2_2 | lAmyg_R | 1.335 | 0.240 | 0.718 | 0.594 | 1.142 | 0.379 |
| 215 | Hipp_L_2_1 | rHipp_L | 1.513 | 0.192 | 1.184 | 0.372 | 0.983 | 0.451 |
| 216 | Hipp_R_2_1 | rHipp_R | -1.448 | 0.206 | -1.779 | 0.177 | -0.513 | 0.701 |
| 217 | Hipp_L_2_2 | cHipp_L | 1.462 | 0.204 | 1.153 | 0.385 | 0.961 | 0.461 |
| 218 | Hipp_R_2_2 | cHipp_R | 2.905 | 0.017 | 2.389 | 0.070 | 1.627 | 0.227 |
| 219 | BG_L_6_1 | vCa_L | 0.425 | 0.721 | 0.169 | 0.893 | 0.459 | 0.725 |
| 220 | BG_R_6_1 | vCa_R | 0.614 | 0.596 | 0.872 | 0.507 | -0.152 | 0.900 |
| 221 | BG_L_6_2 | GP_L | 0.818 | 0.476 | 0.251 | 0.848 | 0.993 | 0.450 |
| 222 | BG_R_6_2 | GP_R | 1.664 | 0.149 | 0.224 | 0.861 | 3.501 | 0.040 |
| 223 | BG_L_6_3 | NAC_L | -0.612 | 0.596 | -0.911 | 0.489 | 0.536 | 0.689 |
| 224 | BG_R_6_3 | NAC_R | -0.337 | 0.783 | -0.706 | 0.600 | 0.490 | 0.711 |
| 225 | BG_L_6_4 | vmPu_L | -0.280 | 0.819 | 0.196 | 0.875 | -0.716 | 0.587 |
| 226 | BG_R_6_4 | vmPu_R | -0.751 | 0.513 | -2.483 | 0.063 | 1.082 | 0.407 |
| 227 | BG_L_6_5 | dCa_L | 1.276 | 0.262 | 1.758 | 0.182 | 0.246 | 0.850 |
| 228 | BG_R_6_5 | dCa_R | 1.462 | 0.204 | 1.721 | 0.191 | 0.696 | 0.594 |
| 229 | BG_L_6_6 | dlPu_L | 1.342 | 0.238 | 1.386 | 0.294 | 0.210 | 0.869 |
| 230 | BG_R_6_6 | dlPu_R | 1.460 | 0.204 | 1.561 | 0.229 | 0.495 | 0.710 |
| 231 | Tha_L_8_1 | mPFtha_L | 0.839 | 0.466 | 0.763 | 0.567 | 0.331 | 0.796 |
| 232 | Tha_R_8_1 | mPFtha_R | 1.149 | 0.310 | 1.829 | 0.168 | -0.721 | 0.587 |
| 233 | Tha_L_8_2 | mPMtha_L | -0.109 | 0.929 | -0.111 | 0.924 | -0.019 | 0.985 |
| 234 | Tha_R_8_2 | mPMtha_R | 1.798 | 0.120 | 1.666 | 0.201 | 0.825 | 0.533 |
| 235 | Tha_L_8_3 | Stha_L | 1.675 | 0.148 | 1.236 | 0.353 | 4.325 | 0.028 |
| 236 | Tha_R_8_3 | Stha_R | 2.099 | 0.070 | 1.673 | 0.200 | 1.520 | 0.253 |
| 237 | Tha_L_8_4 | rTtha_L | 0.241 | 0.838 | -0.868 | 0.507 | 1.786 | 0.196 |
| 238 | Tha_R_8_4 | rTtha_R | 0.593 | 0.607 | 0.630 | 0.642 | 0.178 | 0.891 |
| 239 | Tha_L_8_5 | PPtha_L | 0.211 | 0.855 | 0.505 | 0.706 | -0.373 | 0.773 |
| 240 | Tha_R_8_5 | PPtha_R | -0.525 | 0.652 | 0.965 | 0.464 | -1.506 | 0.257 |
| 241 | Tha_L_8_6 | Otha_L | -0.872 | 0.446 | -0.522 | 0.699 | -0.700 | 0.594 |

| | | | | | | | | |
|---|---|---|---|---|---|---|---|---|
| 242 | Tha_R_8_6 | Otha_R | -0.027 | 0.983 | 0.318 | 0.800 | -0.317 | 0.803 |
| 243 | Tha_L_8_7 | cTtha_L | -0.926 | 0.418 | -0.039 | 0.973 | -1.270 | 0.336 |
| 244 | Tha_R_8_7 | cTtha_R | 1.292 | 0.257 | 1.453 | 0.265 | 0.386 | 0.765 |
| 245 | Tha_L_8_8 | lPFtha_L | 5.082 | 0.000 | 3.657 | 0.015 | 3.822 | 0.030 |
| 246 | Tha_R_8_8 | lPFtha_R | 1.663 | 0.149 | 0.371 | 0.786 | 1.948 | 0.157 |

Note: **a**: *t*-test between all NCs vs. their matched HCs; *P* value was adjusted with FDR correction. **b**: *t*-test between all NC-1s vs. their matched HCs; *P* value was adjusted with FDR correction. **c**: *t*-test between all NC-2s vs. their matched HCs; *P* value was adjusted with FDR correction.

**Supplementary Table 3 Nodal efficiency comparison**

| ROI | Brainnetome region | Subregion name | *t* value[a] (HC vs. NC) | *P*-adj[a] (HC vs. NC) | *t* value[b] (HC vs. NC-1) | *P*-adj[b] (HC vs. NC-1) | *t* value[c] (HC vs. NC-2) | *P*-adj[c] (HC vs. NC-2) |
|---|---|---|---|---|---|---|---|---|
| 1 | SFG_L_7_1 | A8m_L | 2.256 | 0.037 | 2.450 | 0.034 | 0.870 | 0.431 |
| 2 | SFG_R_7_1 | A8m_R | 3.840 | 0.001 | 3.703 | 0.003 | 2.055 | 0.085 |
| 3 | SFG_L_7_2 | A8dl_L | 5.604 | 0.000 | 4.719 | 0.001 | 3.089 | 0.019 |
| 4 | SFG_R_7_2 | A8dl_R | 6.507 | 0.000 | 4.644 | 0.001 | 4.692 | 0.006 |
| 5 | SFG_L_7_3 | A9l_L | 7.075 | 0.000 | 4.862 | 0.001 | 5.236 | 0.004 |
| 6 | SFG_R_7_3 | A9l_R | 7.017 | 0.000 | 4.341 | 0.001 | 5.931 | 0.002 |
| 7 | SFG_L_7_4 | A6dl_L | 3.936 | 0.001 | 2.918 | 0.015 | 2.612 | 0.036 |
| 8 | SFG_R_7_4 | A6dl_R | 3.499 | 0.002 | 3.563 | 0.004 | 1.949 | 0.097 |
| 9 | SFG_L_7_5 | A6m_L | -0.312 | 0.764 | -0.108 | 0.919 | -0.331 | 0.761 |
| 10 | SFG_R_7_5 | A6m_R | 1.786 | 0.094 | 0.987 | 0.372 | 1.515 | 0.186 |
| 11 | SFG_L_7_6 | A9m_L | 6.604 | 0.000 | 4.555 | 0.001 | 4.698 | 0.006 |
| 12 | SFG_R_7_6 | A9m_R | 6.103 | 0.000 | 4.492 | 0.001 | 4.033 | 0.009 |
| 13 | SFG_L_7_7 | A10m_L | 6.372 | 0.000 | 4.561 | 0.001 | 4.553 | 0.007 |
| 14 | SFG_R_7_7 | A10m_R | 5.651 | 0.000 | 2.859 | 0.016 | 6.929 | 0.001 |
| 15 | MFG_L_7_1 | A9/46d_L | 6.712 | 0.000 | 5.452 | 0.000 | 4.314 | 0.009 |
| 16 | MFG_R_7_1 | A9/46d_R | 6.643 | 0.000 | 6.056 | 0.000 | 3.456 | 0.014 |
| 17 | MFG_L_7_2 | IFJ_L | 4.809 | 0.000 | 3.874 | 0.003 | 2.890 | 0.025 |
| 18 | MFG_R_7_2 | IFJ_R | 5.965 | 0.000 | 6.366 | 0.000 | 2.663 | 0.035 |
| 19 | MFG_L_7_3 | A46_L | 5.120 | 0.000 | 5.605 | 0.000 | 2.413 | 0.049 |
| 20 | MFG_R_7_3 | A46_R | 9.716 | 0.000 | 7.236 | 0.000 | 6.231 | 0.002 |
| 21 | MFG_L_7_4 | A9/46v_L | 6.790 | 0.000 | 4.264 | 0.001 | 5.569 | 0.004 |
| 22 | MFG_R_7_4 | A9/46v_R | 7.546 | 0.000 | 6.717 | 0.000 | 4.144 | 0.009 |
| 23 | MFG_L_7_5 | A8vl_L | 6.851 | 0.000 | 5.892 | 0.000 | 3.899 | 0.010 |
| 24 | MFG_R_7_5 | A8vl_R | 9.618 | 0.000 | 8.295 | 0.000 | 5.375 | 0.004 |
| 25 | MFG_L_7_6 | A6vl_L | 5.238 | 0.000 | 3.363 | 0.006 | 4.096 | 0.009 |
| 26 | MFG_R_7_6 | A6vl_R | 6.179 | 0.000 | 5.838 | 0.000 | 3.127 | 0.019 |
| 27 | MFG_L_7_7 | A10l_L | 5.465 | 0.000 | 3.882 | 0.003 | 3.727 | 0.010 |
| 28 | MFG_R_7_7 | A10l_R | 9.402 | 0.000 | 6.394 | 0.000 | 7.376 | 0.001 |
| 29 | IFG_L_6_1 | A44d_L | 4.973 | 0.000 | 3.726 | 0.003 | 3.311 | 0.016 |
| 30 | IFG_R_6_1 | A44d_R | 4.689 | 0.000 | 3.981 | 0.002 | 2.874 | 0.025 |
| 31 | IFG_L_6_2 | IFS_L | 7.355 | 0.000 | 5.802 | 0.000 | 4.486 | 0.007 |
| 32 | IFG_R_6_2 | IFS_R | 5.171 | 0.000 | 3.296 | 0.007 | 4.072 | 0.009 |
| 33 | IFG_L_6_3 | A45c_L | 5.551 | 0.000 | 4.605 | 0.001 | 3.295 | 0.016 |
| 34 | IFG_R_6_3 | A45c_R | 7.830 | 0.000 | 5.831 | 0.000 | 5.021 | 0.005 |
| 35 | IFG_L_6_4 | A45r_L | 4.765 | 0.000 | 4.313 | 0.001 | 2.526 | 0.041 |
| 36 | IFG_R_6_4 | A45r_R | 7.239 | 0.000 | 4.422 | 0.001 | 6.705 | 0.001 |
| 37 | IFG_L_6_5 | A44op_L | 6.996 | 0.000 | 6.100 | 0.000 | 3.951 | 0.010 |
| 38 | IFG_R_6_5 | A44op_R | 6.531 | 0.000 | 6.736 | 0.000 | 3.142 | 0.019 |
| 39 | IFG_L_6_6 | A44v_L | 4.791 | 0.000 | 3.712 | 0.003 | 3.228 | 0.017 |
| 40 | IFG_R_6_6 | A44v_R | 6.865 | 0.000 | 5.700 | 0.000 | 3.974 | 0.010 |
| 41 | OrG_L_6_1 | A14m_L | 5.508 | 0.000 | 3.537 | 0.004 | 4.579 | 0.007 |
| 42 | OrG_R_6_1 | A14m_R | 2.723 | 0.013 | 0.964 | 0.378 | 3.951 | 0.010 |
| 43 | OrG_L_6_2 | A12/47o_L | 5.218 | 0.000 | 3.568 | 0.004 | 3.825 | 0.010 |
| 44 | OrG_R_6_2 | A12/47o_R | 7.385 | 0.000 | 6.940 | 0.000 | 3.794 | 0.010 |
| 45 | OrG_L_6_3 | A11l_L | 4.624 | 0.000 | 3.680 | 0.004 | 3.345 | 0.015 |

| | | | | | | | | |
|---|---|---|---|---|---|---|---|---|
| 46 | OrG_R_6_3 | A11l_R | 5.748 | 0.000 | 4.275 | 0.001 | 3.911 | 0.010 |
| 47 | OrG_L_6_4 | A11m_L | 5.728 | 0.000 | 3.991 | 0.002 | 4.250 | 0.009 |
| 48 | OrG_R_6_4 | A11m_R | 5.285 | 0.000 | 3.996 | 0.002 | 3.344 | 0.015 |
| 49 | OrG_L_6_5 | A13_L | 5.467 | 0.000 | 2.773 | 0.019 | 5.984 | 0.002 |
| 50 | OrG_R_6_5 | A13_R | 4.856 | 0.000 | 4.452 | 0.001 | 2.362 | 0.052 |
| 51 | OrG_L_6_6 | A12/47l_L | 6.737 | 0.000 | 4.795 | 0.001 | 4.641 | 0.007 |
| 52 | OrG_R_6_6 | A12/47l_R | 7.736 | 0.000 | 6.872 | 0.000 | 4.097 | 0.009 |
| 53 | PrG_L_6_1 | A4hf_L | 5.024 | 0.000 | 3.280 | 0.007 | 3.818 | 0.010 |
| 54 | PrG_R_6_1 | A4hf_R | 4.363 | 0.000 | 4.239 | 0.001 | 2.148 | 0.073 |
| 55 | PrG_L_6_2 | A6cdl_L | 5.540 | 0.000 | 5.628 | 0.000 | 3.351 | 0.015 |
| 56 | PrG_R_6_2 | A6cdl_R | 3.363 | 0.003 | 3.654 | 0.004 | 1.454 | 0.204 |
| 57 | PrG_L_6_3 | A4ul_L | 2.126 | 0.048 | 1.124 | 0.308 | 1.838 | 0.116 |
| 58 | PrG_R_6_3 | A4ul_R | 3.385 | 0.003 | 4.244 | 0.001 | 1.610 | 0.160 |
| 59 | PrG_L_6_4 | A4t_L | 0.809 | 0.437 | 0.306 | 0.786 | 0.768 | 0.488 |
| 60 | PrG_R_6_4 | A4t_R | 0.964 | 0.362 | 0.251 | 0.822 | 1.101 | 0.328 |
| 61 | PrG_L_6_5 | A4tl_L | 4.753 | 0.000 | 3.726 | 0.003 | 3.112 | 0.019 |
| 62 | PrG_R_6_5 | A4tl_R | 4.816 | 0.000 | 4.951 | 0.001 | 2.271 | 0.060 |
| 63 | PrG_L_6_6 | A6cvl_L | 5.595 | 0.000 | 4.018 | 0.002 | 3.821 | 0.010 |
| 64 | PrG_R_6_6 | A6cvl_R | 5.881 | 0.000 | 5.043 | 0.001 | 3.743 | 0.010 |
| 65 | PCL_L_2_1 | A1/2/3ll_L | 2.893 | 0.009 | 1.861 | 0.096 | 2.199 | 0.067 |
| 66 | PCL_R_2_1 | A1/2/3ll_R | 0.176 | 0.862 | 0.225 | 0.835 | -0.024 | 0.981 |
| 67 | PCL_L_2_2 | A4ll_L | 2.820 | 0.010 | 2.749 | 0.020 | 1.348 | 0.237 |
| 68 | PCL_R_2_2 | A4ll_R | 2.275 | 0.035 | 1.334 | 0.231 | 1.966 | 0.095 |
| 69 | STG_L_6_1 | A38m_L | 5.355 | 0.000 | 3.862 | 0.003 | 3.661 | 0.011 |
| 70 | STG_R_6_1 | A38m_R | 6.649 | 0.000 | 5.200 | 0.000 | 4.021 | 0.009 |
| 71 | STG_L_6_2 | A41/42_L | 6.322 | 0.000 | 5.054 | 0.001 | 3.870 | 0.010 |
| 72 | STG_R_6_2 | A41/42_R | 4.702 | 0.000 | 3.415 | 0.006 | 3.358 | 0.015 |
| 73 | STG_L_6_3 | TE1.0/TE1.2_L | 3.759 | 0.001 | 2.442 | 0.034 | 2.836 | 0.027 |
| 74 | STG_R_6_3 | TE1.0/TE1.2_R | 5.422 | 0.000 | 5.017 | 0.001 | 2.804 | 0.028 |
| 75 | STG_L_6_4 | A22c_L | 4.216 | 0.000 | 2.369 | 0.039 | 4.138 | 0.009 |
| 76 | STG_R_6_4 | A22c_R | 5.708 | 0.000 | 7.566 | 0.000 | 2.508 | 0.042 |
| 77 | STG_L_6_5 | A38l_L | 3.306 | 0.003 | 2.382 | 0.038 | 2.223 | 0.065 |
| 78 | STG_R_6_5 | A38l_R | 5.193 | 0.000 | 3.433 | 0.005 | 3.938 | 0.010 |
| 79 | STG_L_6_6 | A22r_L | 3.695 | 0.001 | 1.966 | 0.081 | 3.384 | 0.015 |
| 80 | STG_R_6_6 | A22r_R | 4.725 | 0.000 | 3.957 | 0.002 | 2.591 | 0.037 |
| 81 | MTG_L_4_1 | A21c_L | 5.614 | 0.000 | 4.694 | 0.001 | 3.317 | 0.016 |
| 82 | MTG_R_4_1 | A21c_R | 6.209 | 0.000 | 5.329 | 0.000 | 3.859 | 0.010 |
| 83 | MTG_L_4_2 | A21r_L | 4.502 | 0.000 | 3.658 | 0.004 | 2.738 | 0.031 |
| 84 | MTG_R_4_2 | A21r_R | 6.237 | 0.000 | 4.995 | 0.001 | 3.806 | 0.010 |
| 85 | MTG_L_4_3 | A37dl_L | 4.860 | 0.000 | 3.997 | 0.002 | 2.737 | 0.031 |
| 86 | MTG_R_4_3 | A37dl_R | 4.269 | 0.000 | 4.250 | 0.001 | 2.454 | 0.046 |
| 87 | MTG_L_4_4 | aSTS_L | 4.261 | 0.000 | 2.748 | 0.020 | 3.259 | 0.017 |
| 88 | MTG_R_4_4 | aSTS_R | 6.003 | 0.000 | 5.444 | 0.000 | 3.300 | 0.016 |
| 89 | ITG_L_7_1 | A20iv_L | 6.045 | 0.000 | 4.297 | 0.001 | 4.182 | 0.009 |
| 90 | ITG_R_7_1 | A20iv_R | 5.420 | 0.000 | 6.859 | 0.000 | 2.361 | 0.052 |
| 91 | ITG_L_7_2 | A37elv_L | 4.225 | 0.000 | 3.451 | 0.005 | 2.616 | 0.036 |
| 92 | ITG_R_7_2 | A37elv_R | 2.973 | 0.007 | 1.869 | 0.095 | 2.347 | 0.053 |
| 93 | ITG_L_7_3 | A20r_L | 3.961 | 0.001 | 2.558 | 0.028 | 3.142 | 0.019 |
| 94 | ITG_R_7_3 | A20r_R | 5.116 | 0.000 | 5.163 | 0.000 | 2.342 | 0.054 |

| | | | | | | | | |
|---|---|---|---|---|---|---|---|---|
| 95 | ITG_L_7_4 | A20il_L | 4.182 | 0.000 | 2.435 | 0.035 | 3.568 | 0.012 |
| 96 | ITG_R_7_4 | A20il_R | 8.165 | 0.000 | 6.658 | 0.000 | 4.760 | 0.006 |
| 97 | ITG_L_7_5 | A37vl_L | 4.841 | 0.000 | 5.495 | 0.000 | 1.779 | 0.126 |
| 98 | ITG_R_7_5 | A37vl_R | 4.173 | 0.000 | 3.576 | 0.004 | 2.624 | 0.036 |
| 99 | ITG_L_7_6 | A20cl_L | 5.045 | 0.000 | 3.562 | 0.004 | 3.485 | 0.014 |
| 100 | ITG_R_7_6 | A20cl_R | 6.458 | 0.000 | 5.348 | 0.000 | 3.684 | 0.011 |
| 101 | ITG_L_7_7 | A20cv_L | 5.339 | 0.000 | 4.213 | 0.002 | 3.427 | 0.014 |
| 102 | ITG_R_7_7 | A20cv_R | 4.590 | 0.000 | 4.811 | 0.001 | 1.801 | 0.122 |
| 103 | FuG_L_3_1 | A20rv_L | 5.199 | 0.000 | 4.155 | 0.002 | 3.073 | 0.019 |
| 104 | FuG_R_3_1 | A20rv_R | 6.407 | 0.000 | 5.476 | 0.000 | 3.787 | 0.010 |
| 105 | FuG_L_3_2 | A37mv_L | 5.264 | 0.000 | 4.988 | 0.001 | 2.737 | 0.031 |
| 106 | FuG_R_3_2 | A37mv_R | 5.423 | 0.000 | 6.880 | 0.000 | 2.459 | 0.046 |
| 107 | FuG_L_3_3 | A37lv_L | 6.533 | 0.000 | 5.854 | 0.000 | 3.376 | 0.015 |
| 108 | FuG_R_3_3 | A37lv_R | 5.873 | 0.000 | 5.612 | 0.000 | 3.040 | 0.020 |
| 109 | PhG_L_6_1 | A35/36r_L | 3.348 | 0.003 | 2.287 | 0.045 | 2.410 | 0.049 |
| 110 | PhG_R_6_1 | A35/36r_R | 6.579 | 0.000 | 5.218 | 0.000 | 3.899 | 0.010 |
| 111 | PhG_L_6_2 | A35/36c_L | 3.851 | 0.001 | 2.152 | 0.058 | 3.981 | 0.010 |
| 112 | PhG_R_6_2 | A35/36c_R | 3.905 | 0.001 | 2.427 | 0.035 | 3.278 | 0.016 |
| 113 | PhG_L_6_3 | TL_L | 2.882 | 0.009 | 3.134 | 0.010 | 1.002 | 0.369 |
| 114 | PhG_R_6_3 | TL_R | 3.671 | 0.001 | 2.445 | 0.034 | 2.692 | 0.033 |
| 115 | PhG_L_6_4 | A28/34_L | 3.030 | 0.006 | 3.754 | 0.003 | 0.932 | 0.403 |
| 116 | PhG_R_6_4 | A28/34_R | 2.329 | 0.032 | 1.315 | 0.237 | 2.086 | 0.081 |
| 117 | PhG_L_6_5 | TI_L | 1.354 | 0.204 | 0.757 | 0.488 | 1.114 | 0.322 |
| 118 | PhG_R_6_5 | TI_R | 3.633 | 0.001 | 1.918 | 0.088 | 3.433 | 0.014 |
| 119 | PhG_L_6_6 | TH_L | 2.813 | 0.011 | 2.079 | 0.066 | 2.299 | 0.058 |
| 120 | PhG_R_6_6 | TH_R | 4.115 | 0.000 | 2.742 | 0.020 | 3.144 | 0.019 |
| 121 | pSTS_L_2_1 | rpSTS_L | 4.304 | 0.000 | 3.149 | 0.009 | 2.946 | 0.023 |
| 122 | pSTS_R_2_1 | rpSTS_R | 3.376 | 0.003 | 2.987 | 0.013 | 1.658 | 0.152 |
| 123 | pSTS_L_2_2 | cpSTS_L | 5.851 | 0.000 | 4.131 | 0.002 | 4.026 | 0.009 |
| 124 | pSTS_R_2_2 | cpSTS_R | 5.852 | 0.000 | 4.017 | 0.002 | 4.181 | 0.009 |
| 125 | SPL_L_5_1 | A7r_L | 4.101 | 0.000 | 3.742 | 0.003 | 2.397 | 0.050 |
| 126 | SPL_R_5_1 | A7r_R | 5.153 | 0.000 | 4.253 | 0.001 | 3.084 | 0.019 |
| 127 | SPL_L_5_2 | A7c_L | 6.001 | 0.000 | 4.700 | 0.001 | 3.606 | 0.012 |
| 128 | SPL_R_5_2 | A7c_R | 5.095 | 0.000 | 4.196 | 0.002 | 3.090 | 0.019 |
| 129 | SPL_L_5_3 | A5l_L | 3.645 | 0.001 | 3.835 | 0.003 | 1.372 | 0.230 |
| 130 | SPL_R_5_3 | A5l_R | 5.850 | 0.000 | 3.873 | 0.003 | 4.700 | 0.006 |
| 131 | SPL_L_5_4 | A7pc_L | 3.855 | 0.001 | 2.689 | 0.022 | 2.687 | 0.033 |
| 132 | SPL_R_5_4 | A7pc_R | 2.895 | 0.009 | 2.728 | 0.020 | 1.360 | 0.233 |
| 133 | SPL_L_5_5 | A7ip_L | 5.509 | 0.000 | 4.954 | 0.001 | 2.949 | 0.023 |
| 134 | SPL_R_5_5 | A7ip_R | 5.145 | 0.000 | 5.205 | 0.000 | 2.539 | 0.041 |
| 135 | IPL_L_6_1 | A39c_L | 6.408 | 0.000 | 5.834 | 0.000 | 3.228 | 0.017 |
| 136 | IPL_R_6_1 | A39c_R | 6.566 | 0.000 | 9.410 | 0.000 | 2.572 | 0.039 |
| 137 | IPL_L_6_2 | A39rd_L | 5.741 | 0.000 | 5.945 | 0.000 | 2.618 | 0.036 |
| 138 | IPL_R_6_2 | A39rd_R | 6.110 | 0.000 | 4.966 | 0.001 | 3.683 | 0.011 |
| 139 | IPL_L_6_3 | A40rd_L | 6.402 | 0.000 | 4.872 | 0.001 | 4.104 | 0.009 |
| 140 | IPL_R_6_3 | A40rd_R | 5.714 | 0.000 | 5.311 | 0.000 | 3.220 | 0.017 |
| 141 | IPL_L_6_4 | A40c_L | 5.296 | 0.000 | 4.323 | 0.001 | 3.198 | 0.018 |
| 142 | IPL_R_6_4 | A40c_R | 4.961 | 0.000 | 3.838 | 0.003 | 3.400 | 0.015 |
| 143 | IPL_L_6_5 | A39rv_L | 5.405 | 0.000 | 4.914 | 0.001 | 2.936 | 0.023 |

| | | | | | | | | |
|---|---|---|---|---|---|---|---|---|
| 144 | IPL_R_6_5 | A39rv_R | 5.554 | 0.000 | 5.144 | 0.001 | 3.260 | 0.017 |
| 145 | IPL_L_6_6 | A40rv_L | 6.649 | 0.000 | 4.757 | 0.001 | 4.544 | 0.007 |
| 146 | IPL_R_6_6 | A40rv_R | 6.601 | 0.000 | 6.339 | 0.000 | 3.320 | 0.016 |
| 147 | PCun_L_4_1 | A7m_L | 6.033 | 0.000 | 4.621 | 0.001 | 3.736 | 0.010 |
| 148 | PCun_R_4_1 | A7m_R | 3.634 | 0.001 | 3.219 | 0.008 | 1.947 | 0.097 |
| 149 | PCun_L_4_2 | A5m_L | 3.717 | 0.001 | 3.567 | 0.004 | 1.629 | 0.158 |
| 150 | PCun_R_4_2 | A5m_R | 3.452 | 0.002 | 3.781 | 0.003 | 1.093 | 0.329 |
| 151 | PCun_L_4_3 | dmPOS_L | 4.944 | 0.000 | 2.589 | 0.026 | 5.023 | 0.005 |
| 152 | PCun_R_4_3 | dmPOS_R | 6.645 | 0.000 | 4.473 | 0.001 | 4.902 | 0.006 |
| 153 | PCun_L_4_4 | A31_L | 1.678 | 0.115 | 0.331 | 0.770 | 1.836 | 0.116 |
| 154 | PCun_R_4_4 | A31_R | 3.596 | 0.002 | 2.507 | 0.031 | 2.511 | 0.042 |
| 155 | PoG_L_4_1 | A1/2/3ulhf_L | 7.127 | 0.000 | 7.246 | 0.000 | 3.437 | 0.014 |
| 156 | PoG_R_4_1 | A1/2/3ulhf_R | 4.975 | 0.000 | 4.873 | 0.001 | 2.607 | 0.037 |
| 157 | PoG_L_4_2 | A1/2/3tonIa_L | 4.980 | 0.000 | 4.465 | 0.001 | 2.768 | 0.030 |
| 158 | PoG_R_4_2 | A1/2/3tonIa_R | 3.993 | 0.001 | 3.486 | 0.005 | 2.168 | 0.071 |
| 159 | PoG_L_4_3 | A2_L | 4.723 | 0.000 | 3.970 | 0.002 | 3.068 | 0.019 |
| 160 | PoG_R_4_3 | A2_R | 5.482 | 0.000 | 4.580 | 0.001 | 3.110 | 0.019 |
| 161 | PoG_L_4_4 | A1/2/3tru_L | 3.348 | 0.003 | 2.840 | 0.017 | 2.039 | 0.086 |
| 162 | PoG_R_4_4 | A1/2/3tru_R | 3.013 | 0.007 | 2.848 | 0.016 | 1.543 | 0.179 |
| 163 | INS_L_6_1 | G_L | 2.003 | 0.062 | 1.829 | 0.101 | 1.125 | 0.321 |
| 164 | INS_R_6_1 | G_R | 2.871 | 0.009 | 3.586 | 0.004 | 0.608 | 0.583 |
| 165 | INS_L_6_2 | vIa_L | 4.404 | 0.000 | 2.809 | 0.018 | 3.475 | 0.014 |
| 166 | INS_R_6_2 | vIa_R | 6.661 | 0.000 | 5.656 | 0.000 | 3.763 | 0.010 |
| 167 | INS_L_6_3 | dIa_L | 4.935 | 0.000 | 4.759 | 0.001 | 2.866 | 0.025 |
| 168 | INS_R_6_3 | dIa_R | 4.098 | 0.000 | 3.249 | 0.008 | 2.419 | 0.049 |
| 169 | INS_L_6_4 | vId/vIg_L | 3.133 | 0.005 | 2.691 | 0.022 | 1.624 | 0.158 |
| 170 | INS_R_6_4 | vId/vIg_R | 4.892 | 0.000 | 4.418 | 0.001 | 2.611 | 0.036 |
| 171 | INS_L_6_5 | dIg_L | 0.928 | 0.380 | 0.784 | 0.474 | 0.473 | 0.666 |
| 172 | INS_R_6_5 | dIg_R | 2.076 | 0.053 | 1.004 | 0.365 | 2.007 | 0.090 |
| 173 | INS_L_6_6 | dId_L | 3.196 | 0.004 | 2.944 | 0.014 | 1.615 | 0.160 |
| 174 | INS_R_6_6 | dId_R | 2.366 | 0.029 | 1.881 | 0.094 | 1.379 | 0.228 |
| 175 | CG_L_7_1 | A23d_L | 2.636 | 0.016 | 1.801 | 0.106 | 2.046 | 0.086 |
| 176 | CG_R_7_1 | A23d_R | 2.991 | 0.007 | 2.971 | 0.013 | 1.290 | 0.255 |
| 177 | CG_L_7_2 | A24rv_L | 2.276 | 0.035 | 1.827 | 0.101 | 1.343 | 0.237 |
| 178 | CG_R_7_2 | A24rv_R | 0.999 | 0.346 | 0.420 | 0.706 | 1.171 | 0.301 |
| 179 | CG_L_7_3 | A32p_L | 6.185 | 0.000 | 4.041 | 0.002 | 4.912 | 0.006 |
| 180 | CG_R_7_3 | A32p_R | 3.133 | 0.005 | 2.710 | 0.021 | 1.718 | 0.138 |
| 181 | CG_L_7_4 | A23v_L | 1.069 | 0.314 | 0.614 | 0.574 | 0.889 | 0.423 |
| 182 | CG_R_7_4 | A23v_R | 5.402 | 0.000 | 4.630 | 0.001 | 2.881 | 0.025 |
| 183 | CG_L_7_5 | A24cd_L | 4.054 | 0.001 | 3.527 | 0.005 | 2.218 | 0.066 |
| 184 | CG_R_7_5 | A24cd_R | 3.446 | 0.002 | 3.625 | 0.004 | 1.118 | 0.322 |
| 185 | CG_L_7_6 | A23c_L | 0.896 | 0.396 | 1.275 | 0.251 | 0.060 | 0.957 |
| 186 | CG_R_7_6 | A23c_R | 0.285 | 0.781 | 1.247 | 0.260 | -0.692 | 0.532 |
| 187 | CG_L_7_7 | A32sg_L | 3.164 | 0.005 | 1.202 | 0.276 | 3.842 | 0.010 |
| 188 | CG_R_7_7 | A32sg_R | 5.219 | 0.000 | 4.369 | 0.001 | 3.213 | 0.017 |
| 189 | MVOcC_L_5_1 | cLinG_L | 5.125 | 0.000 | 3.927 | 0.002 | 3.193 | 0.018 |
| 190 | MVOcC_R_5_1 | cLinG_R | 5.018 | 0.000 | 4.182 | 0.002 | 2.900 | 0.025 |
| 191 | MVOcC_L_5_2 | rCunG_L | 6.153 | 0.000 | 4.123 | 0.002 | 4.554 | 0.007 |
| 192 | MVOcC_R_5_2 | rCunG_R | 4.520 | 0.000 | 3.150 | 0.009 | 3.184 | 0.018 |

| | | | | | | | | |
|---|---|---|---|---|---|---|---|---|
| 193 | MVOcC_L_5_3 | cCunG_L | 5.924 | 0.000 | 4.149 | 0.002 | 4.138 | 0.009 |
| 194 | MVOcC_R_5_3 | cCunG_R | 6.430 | 0.000 | 5.201 | 0.000 | 3.775 | 0.010 |
| 195 | MVOcC_L_5_4 | rLinG_L | 5.451 | 0.000 | 3.572 | 0.004 | 4.152 | 0.009 |
| 196 | MVOcC_R_5_4 | rLinG_R | 4.443 | 0.000 | 3.866 | 0.003 | 3.066 | 0.019 |
| 197 | MVOcC_L_5_5 | vmPOS_L | 6.344 | 0.000 | 3.398 | 0.006 | 7.193 | 0.001 |
| 198 | MVOcC_R_5_5 | vmPOS_R | 4.343 | 0.000 | 2.569 | 0.027 | 3.661 | 0.011 |
| 199 | LOcC_L_4_1 | mOccG_L | 6.738 | 0.000 | 6.031 | 0.000 | 3.597 | 0.012 |
| 200 | LOcC_R_4_1 | mOccG_R | 6.657 | 0.000 | 4.243 | 0.001 | 5.326 | 0.004 |
| 201 | LOcC_L_4_2 | V5/MT+_L | 5.971 | 0.000 | 5.545 | 0.000 | 3.013 | 0.021 |
| 202 | LOcC_R_4_2 | V5/MT+_R | 7.035 | 0.000 | 8.619 | 0.000 | 3.466 | 0.014 |
| 203 | LOcC_L_4_3 | OPC_L | 5.684 | 0.000 | 4.437 | 0.001 | 3.448 | 0.014 |
| 204 | LOcC_R_4_3 | OPC_R | 5.676 | 0.000 | 4.284 | 0.001 | 3.665 | 0.011 |
| 205 | LOcC_L_4_4 | iOccG_L | 6.182 | 0.000 | 5.015 | 0.001 | 3.771 | 0.010 |
| 206 | LOcC_R_4_4 | iOccG_R | 6.374 | 0.000 | 4.896 | 0.001 | 4.244 | 0.009 |
| 207 | LOcC_L_2_1 | msOccG_L | 6.532 | 0.000 | 4.887 | 0.001 | 4.166 | 0.009 |
| 208 | LOcC_R_2_1 | msOccG_R | 5.756 | 0.000 | 4.609 | 0.001 | 3.361 | 0.015 |
| 209 | LOcC_L_2_2 | lsOccG_L | 6.006 | 0.000 | 5.171 | 0.000 | 3.227 | 0.017 |
| 210 | LOcC_R_2_2 | lsOccG_R | 6.325 | 0.000 | 4.771 | 0.001 | 4.082 | 0.009 |
| 211 | Amyg_L_2_1 | mAmyg_L | 4.129 | 0.000 | 2.235 | 0.050 | 3.866 | 0.010 |
| 212 | Amyg_R_2_1 | mAmyg_R | 4.780 | 0.000 | 4.270 | 0.001 | 2.854 | 0.026 |
| 213 | Amyg_L_2_2 | lAmyg_L | 1.509 | 0.157 | 0.137 | 0.900 | 1.993 | 0.091 |
| 214 | Amyg_R_2_2 | lAmyg_R | 1.912 | 0.074 | 2.344 | 0.041 | 0.571 | 0.602 |
| 215 | Hipp_L_2_1 | rHipp_L | 3.490 | 0.002 | 3.553 | 0.004 | 1.959 | 0.096 |
| 216 | Hipp_R_2_1 | rHipp_R | 3.647 | 0.001 | 3.173 | 0.009 | 2.112 | 0.077 |
| 217 | Hipp_L_2_2 | cHipp_L | 4.105 | 0.000 | 2.857 | 0.016 | 3.105 | 0.019 |
| 218 | Hipp_R_2_2 | cHipp_R | 5.431 | 0.000 | 4.663 | 0.001 | 3.108 | 0.019 |
| 219 | BG_L_6_1 | vCa_L | 1.268 | 0.235 | 0.797 | 0.467 | 0.973 | 0.383 |
| 220 | BG_R_6_1 | vCa_R | 2.176 | 0.043 | 2.184 | 0.055 | 1.039 | 0.353 |
| 221 | BG_L_6_2 | GP_L | 3.927 | 0.001 | 2.549 | 0.028 | 2.973 | 0.022 |
| 222 | BG_R_6_2 | GP_R | 4.794 | 0.000 | 3.839 | 0.003 | 3.134 | 0.019 |
| 223 | BG_L_6_3 | NAC_L | 1.102 | 0.300 | 0.720 | 0.509 | 0.810 | 0.464 |
| 224 | BG_R_6_3 | NAC_R | 1.177 | 0.270 | 1.233 | 0.264 | 0.515 | 0.639 |
| 225 | BG_L_6_4 | vmPu_L | 1.204 | 0.259 | 0.514 | 0.640 | 1.190 | 0.294 |
| 226 | BG_R_6_4 | vmPu_R | 0.561 | 0.589 | -0.084 | 0.934 | 0.894 | 0.422 |
| 227 | BG_L_6_5 | dCa_L | 4.518 | 0.000 | 3.874 | 0.003 | 2.469 | 0.045 |
| 228 | BG_R_6_5 | dCa_R | 3.963 | 0.001 | 4.337 | 0.001 | 2.005 | 0.090 |
| 229 | BG_L_6_6 | dlPu_L | 5.531 | 0.000 | 4.133 | 0.002 | 3.647 | 0.011 |
| 230 | BG_R_6_6 | dlPu_R | 4.774 | 0.000 | 5.405 | 0.000 | 2.024 | 0.088 |
| 231 | Tha_L_8_1 | mPFtha_L | 0.594 | 0.569 | 0.967 | 0.378 | -0.192 | 0.862 |
| 232 | Tha_R_8_1 | mPFtha_R | 0.828 | 0.428 | 1.599 | 0.150 | -0.607 | 0.583 |
| 233 | Tha_L_8_2 | mPMtha_L | 1.824 | 0.088 | 0.956 | 0.381 | 1.775 | 0.126 |
| 234 | Tha_R_8_2 | mPMtha_R | 5.338 | 0.000 | 4.974 | 0.001 | 2.636 | 0.036 |
| 235 | Tha_L_8_3 | Stha_L | 2.576 | 0.018 | 1.554 | 0.162 | 3.153 | 0.019 |
| 236 | Tha_R_8_3 | Stha_R | 3.316 | 0.003 | 2.985 | 0.013 | 1.643 | 0.155 |
| 237 | Tha_L_8_4 | rTtha_L | 0.873 | 0.407 | -0.232 | 0.834 | 1.283 | 0.257 |
| 238 | Tha_R_8_4 | rTtha_R | 0.629 | 0.548 | 0.812 | 0.461 | 0.068 | 0.955 |
| 239 | Tha_L_8_5 | PPtha_L | 2.317 | 0.033 | 2.146 | 0.058 | 1.015 | 0.364 |
| 240 | Tha_R_8_5 | PPtha_R | 0.536 | 0.603 | 1.005 | 0.365 | -0.346 | 0.753 |
| 241 | Tha_L_8_6 | Otha_L | 1.032 | 0.330 | 1.278 | 0.251 | 0.207 | 0.853 |

| | | | | | | | | |
|---|---|---|---|---|---|---|---|---|
| 242 | Tha_R_8_6 | Otha_R | 0.852 | 0.416 | 0.706 | 0.516 | 0.463 | 0.671 |
| 243 | Tha_L_8_7 | cTtha_L | 1.118 | 0.295 | 0.969 | 0.378 | 0.603 | 0.584 |
| 244 | Tha_R_8_7 | cTtha_R | 2.763 | 0.012 | 2.014 | 0.074 | 1.825 | 0.118 |
| 245 | Tha_L_8_8 | lPFtha_L | 9.920 | 0.000 | 8.819 | 0.000 | 5.376 | 0.004 |
| 246 | Tha_R_8_8 | lPFtha_R | 2.191 | 0.042 | 0.278 | 0.805 | 3.097 | 0.019 |

Note: **a**: *t*-test between all NCs vs. their matched HCs; *P* value was adjusted with FDR correction. **b**: *t*-test between all NC-1s vs. their matched HCs; *P* value was adjusted with FDR correction. **c**: *t*-test between all NC-2s vs. their matched HCs; *P* value was adjusted with FDR correction.

**Supplementary Table 4 Top 20 Panther pathway terms for narcolepsy featured alterations**

| Gene Set | Description | Size | Leading Edge Number | ES | *NES* | *P* value | *FDR q* value |
|---|---|---|---|---|---|---|---|
| P00041 | Metabotropic glutamate receptor group I pathway | 23 | 14 | 0.56838 | 1.9319 | <2.2e-16 | 0.033418 |
| P00037 | Ionotropic glutamate receptor pathway | 43 | 17 | 0.47296 | 1.8986 | <2.2e-16 | 0.029385 |
| P00035 | Interferon-gamma signaling pathway | 23 | 14 | 0.55092 | 1.8927 | 0.0016722 | 0.020358 |
| P00039 | Metabotropic glutamate receptor group III pathway | 61 | 24 | 0.40874 | 1.7458 | <2.2e-16 | 0.056695 |
| P00046 | Oxidative stress response | 42 | 21 | 0.43504 | 1.7333 | 0.0031797 | 0.052047 |
| P00003 | Alzheimer disease-amyloid secretase pathway | 58 | 24 | 0.39232 | 1.6897 | 0.0044843 | 0.068152 |
| P00026 | Heterotrimeric G-protein signaling pathway-Gi alpha and Gs alpha mediated pathway | 128 | 38 | 0.33157 | 1.6351 | 0.0013755 | 0.085145 |
| P00029 | Huntington disease | 119 | 48 | 0.30585 | 1.5065 | 0.011252 | 0.19045 |
| P00021 | FGF signaling pathway | 93 | 36 | 0.32045 | 1.4996 | 0.021067 | 0.18236 |
| P06959 | CCKR signaling map | 154 | 55 | 0.28232 | 1.4445 | 0.017264 | 0.18864 |
| P00005 | Angiogenesis | 141 | 47 | -0.19419 | -1.0996 | 0.22308 | 0.56941 |
| P00034 | Integrin signalling pathway | 148 | 50 | -0.19440 | -1.1133 | 0.21401 | 0.58400 |
| P00052 | TGF-beta signaling pathway | 74 | 24 | -0.22482 | -1.1514 | 0.19932 | 0.67351 |
| P00006 | Apoptosis signaling pathway | 99 | 35 | -0.22025 | -1.1804 | 0.12892 | 0.78527 |
| P04372 | 5-Hydroxytryptamine degredation | 15 | 8 | -0.54220 | -1.7639 | 0.0069930 | 0.12905 |

**Supplementary Table 5 Top 20 Gene Ontology terms of biological process for narcolepsy featured alterations**

| Gene Set | Description | Size | Leading Edge Number | ES | *NES* | *P* value | *FDR q* value |
|---|---|---|---|---|---|---|---|
| GO:0099504 | synaptic vesicle cycle | 180 | 76 | 0.43884 | 2.2901 | <2.2e-16 | <2.2e-16 |
| GO:0050803 | regulation of synapse structure or activity | 206 | 82 | 0.39042 | 2.0691 | <2.2e-16 | 0.0038049 |
| GO:0007215 | glutamate receptor signaling pathway | 85 | 47 | 0.43866 | 2.0484 | <2.2e-16 | 0.0044991 |
| GO:0042391 | regulation of membrane potential | 355 | 151 | 0.35348 | 1.9835 | <2.2e-16 | 0.0072705 |
| GO:0051668 | localization within membrane | 138 | 61 | 0.38549 | 1.9375 | <2.2e-16 | 0.0087410 |
| GO:0007588 | excretion | 48 | 25 | -0.44939 | -2.0553 | <2.2e-16 | 0.019456 |
| GO:0097164 | ammonium ion metabolic process | 160 | 60 | -0.33883 | -1.9779 | <2.2e-16 | 0.021994 |
| GO:0034340 | response to type I interferon | 64 | 27 | -0.39795 | -1.9536 | <2.2e-16 | 0.026505 |
| GO:0048645 | animal organ formation | 43 | 18 | -0.43738 | -1.9146 | <2.2e-16 | 0.032452 |
| GO:0050900 | leukocyte migration | 286 | 122 | -0.30397 | -1.9017 | <2.2e-16 | 0.034259 |
| GO:0007229 | integrin-mediated signaling pathway | 82 | 34 | -0.35747 | -1.8899 | <2.2e-16 | 0.034877 |
| GO:1901342 | regulation of vasculature development | 245 | 93 | -0.28377 | -1.7718 | <2.2e-16 | 0.038436 |
| GO:0051181 | cofactor transport | 37 | 21 | -0.41177 | -1.7476 | 0.0057143 | 0.042875 |
| GO:0032609 | interferon-gamma production | 64 | 29 | -0.36544 | -1.7859 | <2.2e-16 | 0.043467 |
| GO:0051705 | multi-organism behavior | 61 | 26 | 0.39452 | 1.7312 | 0.0030628 | 0.044781 |
| GO:0034067 | protein localization to Golgi apparatus | 24 | 12 | 0.48467 | 1.7154 | 0.0033841 | 0.046257 |
| GO:0072512 | trivalent inorganic cation transport | 32 | 14 | 0.45538 | 1.7199 | 0.0082919 | 0.047827 |

**Supplementary Table 6 Top 10 Gene Ontology terms of cellular components for narcolepsy featured alterations**

| Gene Set | Description | Size | Leading Edge Number | ES | *NES* | *P* value | *FDR q* value |
|---|---|---|---|---|---|---|---|
| GO:0097060 | synaptic membrane | 389 | 195 | 0.40471 | 2.3023 | <2.2e-16 | <2.2e-16 |
| GO:0098978 | glutamatergic synapse | 338 | 153 | 0.39725 | 2.2243 | <2.2e-16 | <2.2e-16 |
| GO:0030667 | secretory granule membrane | 216 | 102 | -0.31657 | -1.9483 | <2.2e-16 | 0.017131 |
| GO:0044455 | mitochondrial membrane part | 206 | 102 | 0.31907 | 1.6930 | <2.2e-16 | 0.028539 |
| GO:0000151 | ubiquitin ligase complex | 256 | 105 | 0.30576 | 1.6685 | <2.2e-16 | 0.034156 |
| GO:0043025 | neuronal cell body | 436 | 106 | 0.28138 | 1.6106 | <2.2e-16 | 0.048628 |
| GO:0030055 | cell-substrate junction | 377 | 110 | -0.24758 | -1.6277 | <2.2e-16 | 0.062268 |
| GO:0098552 | side of membrane | 363 | 138 | -0.24851 | -1.6290 | <2.2e-16 | 0.068981 |
| GO:0031012 | extracellular matrix | 346 | 106 | -0.23960 | -1.5600 | <2.2e-16 | 0.075252 |
| GO:0045177 | apical part of cell | 266 | 70 | -0.22763 | -1.4249 | <2.2e-16 | 0.12359 |
